# Association of Dietary Flavonoid Intake with Incident Depression Risk and Brain Structural Changes: A Prospective Study in the UK Biobank

**DOI:** 10.1101/2025.11.07.25339744

**Authors:** Yaying Cao, Qinhao Guan, Xuewei Yang, Hui Chen, Tianzhu Li, Yuze Li, Zhe Huang, Alejandro Cifuentes, Elena Ibañez, Changzheng Yuan, Weihong Lu

**Author notes:** contributed equally as corresponding authors (Yaying Cao, Changzheng Yuan, Weihong Lu).

## Abstract

**Objectives:** To prospectively evaluate the association of dietary flavonoid intake with incident depression risk and brain structural changes.

**Design:** Prospective cohort study.

**Setting:** UK Biobank.

**Participants:** 114 848 non-depressed individuals (with ≥ 2 eligible 24-h dietary recalls) for depression risk and a subgroup of 2120 for brain structural changes.

**Exposures:** Total flavonoid, the Flavodiet Score (reflecting servings of top 10 flavonoid-rich foods), and specific flavonoid subclasses and individual compounds.

**Main outcome measures:** Incident depression (ascertained via inpatient and death register data) and alterations in cortical volumes and white matter integrity.

**Results:** During a median follow-up of 10.5 years, 2965 incident depression cases were identified. After adjusting for demographic, major lifestyle, and clinical factors, total flavonoid intake and the Flavodiet Score were both linearly associated with a reduced depression risk, with hazard ratios (HRs) and 95% confidence intervals (CIs) for quintile 5 vs. corresponding quintile 1 being 0.84 (0.75 to 0.95) and 0.77 (0.68 to 0.87). No significant interactions were found between total flavonoid intake and potential effect modifiers, such as demographic, lifestyle, and chronic disease factors. Subclass analysis showed that intakes of anthocyanins and flavanones both were linearly associated with a lower depression risk, with HRs (95% CIs) for quintile 5 vs. corresponding quintile being 0.63 (0.55 to 0.71) and 0.84 (0.95 to 0.94). Beyond specific compounds from flavanones and anthocyanins subclasses, proanthocyanidin dimers, catechin, and epicatechin as a cluster derived by the k-means method were also associated with a reduced depression risk. Path diagrams suggested a sequential chain of mediation via sarcopenia biomarkers (creatinine to cystatin C ratio and muscle quality index) and subsequent diabetes status in the association of total flavonoid intake with depression risk. Regarding brain structural changes, higher flavonoid intake was associated with less atrophy in specific cortical regions (left Caudal anterior cingulate, left Supramarginal, right Caudal middle frontal, and right Precentral) and a smaller increase in mean diffusivity in the left Anterior Thalamic Radiation tract (all *P* < 0.05).

**Conclusions:** Our findings suggest that dietary flavonoids, notably anthocyanins and flavanones, may contribute favourably to depression risk reduction and may offer protection against adverse brain structural changes. Further mechanistic studies and randomized controlled trials are warranted to validate these effects and translate them into public health policy.

**Research summary:** *WHAT IS ALREADY KNOWN ON THIS TOPIC:* Flavonoids are known to exert beneficial effects through key mechanisms highly relevant to depression pathophysiology, as shown in preliminary research. Population-based research between flavonoid intake and risk of depression is generally scarce and largely supports the positive impact of total flavonoid intake on depression risk. Nevertheless, the association of flavonoid with depression risk is inconsistent in the subclass level and exploration analysis in the individual compound level is conducted ignoring the correlations in between.

*WHAT THIS STUDY ADDS:* Partial monomers (i.e., catechin and epicatechin) and dimers of proanthocyanidins as a cluster from the k-means method were associated with a reduced depression risk, alongside specific compounds from flavanones and anthocyanins. The effect of total flavonoid intake on depression risk was mediated by a sequential chain of intermediate factors, specifically sarcopenia biomarkers and subsequent diabetes status. The total flavonoid intake and sum of servings for top 10 flavonoids-rich food items presented protection against depression-related brain structural changes.

## Introduction

Major depressive disorder is a leading cause of disability globally, affecting an estimated 5.7% of adults ^[1]^. A drastic deterioration in psychosocial abilities and a marked decline in life quality is experienced by individuals with this condition ^[2]^, accounting for the largest proportion (37.3%) of disability-adjusted life years attributable to mental disorders ^[3]^. The importance of healthy diets in mitigating depression risk is increasingly highlighted by the literature. A reduced risk of depression has been shown to be associated with dietary patterns characterized by low ultra-processed food intake or high nutrient density in observational evidence, and moderate-to-large symptom relief has been demonstrated by randomized controlled trials focusing on the Mediterranean diet ^[4]^. Flavonoids, a class of plant secondary metabolites, are considered to exert beneficial effects through key mechanisms highly relevant to depression pathophysiology, including antioxidant ^[5]^ and anti-inflammation actions ^[6]^, as well as inhibition of monoamine oxidase A ^[7]^ and regulation of the hypothalamic-pituitary-adrenal axis ^[8]^.

Population-based evidence investigating the association between flavonoids and depression risk is generally scarce. A cross-sectional study from the National Health and Nutrition Examination Survey (NHANES) suggested that total flavonoid, along with subclasses as flavanones and flavones, was associated with a lower likelihood of depression among 8183 adults ^[9]^. The only available prospective study was conducted among females from the Nurses’ Health Study (NHS) and NHS II cohorts ^[10]^. This research indicated that total flavonoid intake was associated with a reduced risk of incident depression, a finding observed exclusively in the NHS cohort, a group with an older age profile compared to NHS II. For individual subclasses, inverse associations with incident depression risk were observed for flavonols, flavones, and flavanones based on the pooled data from both cohorts. These studies displayed certain discrepancies in subclass-level associations with depression risk. Regarding potential underlying biological mechanisms of flavonoid intake, cross-sectional analyses from NHANES suggested mediation effects of 13.8% for sleep duration on depression risk ^[9]^ and 9.8% for HDL-c on perimenopausal depression ^[11]^, respectively. While preliminary animal research suggested that specific flavonoid compounds may alleviate total brain atrophy, no population-based studies have examined the specific influence of flavonoid intake on depression-related structural changes in human brain regions.

Leveraging the large-scale, multidimensional prospective data from the UK Biobank (UKB), the effect of dietary flavonoid intake on incident depression risk was investigated across hierarchical levels, including total intake, subclasses, and individual compounds. Specifically, effect modification was explored in various subpopulations and the potential mediating role of a comprehensive set of variables was analyzed. Furthermore, the relationship between flavonoid intake and longitudinal changes in total and cortical brain volume was examined.

## Methods

### Study Population

The UK Biobank project is a large-scale, on-going population-based prospective study with extensive data in phenotypic and multi-omics data collected from over 500,000 individuals aged 40-69 years at the time of recruitment between 2006 and 2010 ^[12]^. This study adheres to the Strengthening the Reporting of Observational Studies in Epidemiology (STROBE) reporting guideline. For the current study, participants were included if they were ≥ 40 years at baseline with appropriate dietary data defined as having ≥ 2 dietary assessments with plausible energy intakes (800-4200 kcal/d for males; 600-3500 kcal/d for females). Participants with a diagnosis or self-report of depression, or a baseline PHQ-2 score of ≥ 3 prior to the second valid dietary assessment were excluded (**Supplementary Figure 1**).

### Flavonoid Intake

Between April 2009 and June 2012, 210 965 individuals in the cohort completed up to five Oxford WebQ 24-h dietary recall assessments, covering 206 food items and 32 beverages consumed over the preceding 24 hours. Accumulated average dietary intake of flavonoids and energy for assessments of dietary recalls with plausible energy intake before the end of follow-up was calculated to reduce within-subject variation and best represent a long-term diet. ^[13]^ A database for the assessment of various flavonoid subclass intakes was developed utilizing food composition data from the US Department of Agriculture (USDA) database and Phenol-Explorer 3.6 (http://phenol-explorer.eu/) as main sources. Flavonoid values were assigned to each food and beverage item listed in the Oxford WebQ. For composite recipes, flavonoid values were derived by assigning values to each ingredient of a standard recipe and subsequently summing the values for the total food. For broadly described foods that lacked specific matching items in the composition database, proportions of relevant items were determined according to nutritional epidemiological surveys, marketing share reports, and official data from the United Kingdom or other European countries. For example, to estimate the compound content of red wines, three NDB numbers (14098, 14097, and 14100) were selected based on Bruwer et al.’s statistics regarding grape varieties prevalent in the UK market ^[14]^. Details were described in **Supplementary Method 1**.

We focused on the following 7 subclasses, which are commonly consumed in the Western diet: flavanones (eriodictyol, hesperetin, and naringenin), anthocyanins (cyanidin, delphinidin, malvidin, pelargonidin, petunidin, and peonidin), flavan-3-ols (catechins and epicatechins), flavonols (quercetin, kaempferol, myricetin, and isohamnetin), flavones (luteolin and apigenin), polymers (including proanthocyanidins [excluding monomers], theaflavins, and thearubigins), and proanthocyanidins separately (dimers, trimers, 4–6mers, 7–10mers, polymers, and monomers). Total flavonoid intake was calculated by summing the 6 component subclasses (flavanones, anthocyanins, flavan-3-ols, flavonols, flavones, and polymers). To further enhance the clinical and practical utility of the study, a flavodiet score was derived by summing the intakes as servings per day of the key contributors to each flavonoid subclass, including tea (black and green), red wine, apples, berries, grapes, oranges, grapefruit, sweet peppers, onions, and dark chocolate as previously described ^[15]^.

### Diagnoses of depression and baseline chronic health conditions

Depression diagnosis was ascertained through baseline self-report interviewed by a trained nurse and then examined by a doctor, baseline PHQ-2 score, hospital inpatient records and linkage to death register data using the International Classification of Disease coding system. A PHQ-2 score of 3 or more was defined as depression. For our analysis, the end of the follow-up period was 31 October 2022 for England, 31 August 2022 for Scotland, and 31 May 2022 for Wales, or the onset of incident depression event, whichever came first. Baseline chronic health conditions considered in this study included myocardial infarction, stroke, breast cancer, prostate cancer, colorectal cancer, vertebrae fracture, hip fracture and dementia. A history of these conditions was defined by their presence at the time of the last 24-h dietary recall. The presence of a specific condition was confirmed if indicated in any of the following sources: baseline self-report, hospital inpatient records, death register, or cancer registry data (**Supplementary Table 1**).

### Brain Structure Assessment

Since 2014, two large-scale magnetic resonance imaging (MRI) assessments of the heads have been conducted among over 40,000 individuals, all acquired by a standard Siemens Skyra 3T scanner with 32-channel head coil. Further details are available at https://www.fmrib.ox.ac.uk/ukbiobank/protocol/ ^[16]^. For the present study, data derived from T1 structural imaging (T1-MRI), T2-weighted brain MRI (T2-MRI), and diffusion-weighted imaging (dMRI) were utilized, which were generated by an image-processing pipeline developed and run on behalf of UK Biobank ^[17]^. Volumes of total grey matter, total white matter, total brain (sum of total grey matter and total white matter) were from T1 images (category 110) and white matter hyperintensities was from T2 images (category 112). Volumes of cortical regions (category 192) were generated using Freesurfer software from T1 images. Microstructural phenotypes of white matter, including the anisotropy (FA) and mean diffusivity (MD) of the white matter tracts (category 135), were generated from dMRI data after correction for eddy currents and head movements. In total, data from 2120 individuals with two brain MRI scans were used. All brain structure indices were normalized to the standard space. The *z* score of each brain structural volumetric marker was calculated as the original value minus the population mean divided by the standard deviation (SD) at the Phase 1 MRI assessment. To represent the changes in these markers, we first calculated the z score of the values from Phase 2 using the population mean and SD at the first MRI assessment and subtracted Phase 2 *z* scores from Phase 1 *z* scores ^[18]^.

### Covariate Definition

Demographic characteristics, lifestyle, depression history, chronic disease and dietary factors were adjusted in multivariable models, including age at last dietrecall, sex (male/female), socioeconomic status (Townsend deprivation index categorized as low [quintile 1], moderate [quintiles 2-4], or high [quintile 5] deprivation), education level (lower secondary, higher secondary, vocational, higher, or other), race (White, Asian, Black, Multiracial, Chinese, or other), smoking history (ever/never), sleep duration (short, moderate, or long), physical activity (regular/irregular), body mass index categories (thin, normal, overweight, or obese), family history of depression (yes/no), regular medications taking (yes/no), number of chronic conditions (none/yes), number of 24-h diet recall assessments (2, 3, 4, or 5), healthy plant-based diet index (as quartiles), alcohol intake from non-red drinks in g per day (low, high, or missing), energy intake (as continuous sex-specific quintiles), and fat intake (as continuous sex-specific quintiles) (Details in **Supplementary Method 2**).

### Statistical Analysis

For variables with a missing rate below 5%, missing data were imputed as medians for continuous variables and as modes for categorical variables; otherwise, a missing category was created (missing rate in **Supplementary Table 2**). Baseline characteristics were presented as mean (SD) for continuous variables and n (%) for categorical variables. Pearson’s First Coefficient (PFC) was calculated as the index of distribution skewness. Hierarchical flavonoid intakes with a PFC >1 were log_2_-transformed, while those with a PFC < −1 were Yeo-johnson transformed (histogram of intakes of total flavonoid, subclasses, and compounds in **Supplementary Figure 2 & 3**). The proportional hazard assumption of Cox proportional hazards model was assessed via the Schoenfeld residual test and was not violated for total flavonoid intake or the Flavodiet Score. A k-means clustering classification of the entire study population was subsequently performed to define flavonoid compound clusters with similar co-exposure profiles, with the optimal number of clusters determined using the Silhouette Coefficient. The Cox proportional hazards model, using age as the time-scale, was applied to examine the association of the Flavodiet Score, intakes of total flavonoid, subclasses, and clusters with incident depression risk. Several sensitivity analyses were conducted using the same models, including those restricted to: (1) participants aged ≥ 60 years at baseline; (2) participants diagnosed with depression within 2 years of follow-up excluded; (3) participants with at least 5 years of follow-up; (4) participants residing in areas of high socioeconomic deprivation or those with low education attainment. Linear regression was applied for analyses of brain structural changes. Additionally, the proportional contribution of a series indices (physical measures, blood biochemistry, and history of diseases) to the association between total flavonoid intake and depression risk was evaluated using causal mediation analysis via the ‘mediation’ package in R. Path analysis using Structural Equation Modeling tested the hypothesized relationships for intermediate factors potentially in a sequential chain and WLSMV and theta parameterization in the R package ‘lavaan’ were utilized for categorical outcomes. Interaction analysis was performed by adding the cross-product of total flavonoid intake with age group (< 60 years or ≥ 60 years) and other covariates, respectively. Significance was tested using maximum likelihood ratio test. All analyses were conducted using R (version 4.4.3), and a two-tailed p<0.05 was considered statistically significant.

### Patient and public involvement

No dedicated funding was available for patient and public involvement in this study. While the UK Biobank resource incorporated extensive prior public consultation in its initial design, patients were not directly involved in key stages of the current project. Specifically, the did not contribute to defining the research questions, setting the outcome measures, developing the study design or implementation plans, nor were they consulted for the interpretation or reporting of the results.

## Results

### Baseline characteristics

A total of 114 848 participants, with a median age of 60.4 years at baseline, were included in the analysis. Participant characteristics stratified by total flavonoid intake quintiles were presented in **Table 1**. Briefly, participants with a higher intake of total flavonoid tended to have higher energy and fat intakes, a higher hPDI score, and were generally older. They were also more likely to be White, never smokers, from a higher socioeconomic status, have moderate sleep duration, and to engage in regular exercises. Baseline characteristics and dietary intakes according to incident depression diagnosis were shown in **Supplementary Table 3**.

**Table 1.** Dietary intake and baseline characteristics by quintiles of total flavonoid intake.

| Characteristics | Value to median (IQR) |  |  |  |  |
| --- | --- | --- | --- | --- | --- |
|  | Quintile 1 (lowest)<br>(n = 22 970) | Quintile 2<br>(n = 22 969) | Quintile 3<br>(n = 22 970) | Quintile 4<br>(n = 22 969) | Quintile 5 (highest)<br>(n = 22 970) |
| **Dietary** |  |  |  |  |  |
| Total flavonoid to mg/d | 194.4 (108.2 to 276.0) | 501.5 (429.1 to 572.1) | 760.5 (700.4 to 822.4) | 985.4 (937.1 to 1033.2) | 1237.0 (1150.4 to 1375.5) |
| Anthocyanins to mg/d | 4.2 (0.9 to 30.6) | 15.5 (1.9 to 96.4) | 17.3 (2.0 to 107.6) | 11.8 (1.7 to 83.8) | 81.0 (9.7 to 190.0) |
| Flavan-3-ols to mg/d | 28.4 (15.3 to 46.3) | 95.2 (69.4 to 120.0) | 154.4 (131.6 to 175.9) | 210.5 (185.9 to 225.4) | 238.8 (220.9 to 257.4) |
| Flavones to mg/d | 0.2 (0.1 to 0.6) | 0.3 (0.1 to 0.7) | 0.3 (0.1 to 0.7) | 0.3 (0.1 to 0.7) | 0.4 (0.2 to 0.8) |
| Flavonols to mg/d | 8.3 (5.0 to 12.3) | 17.7 (13.8 to 21.7) | 26.2 (22.5 to 30.1) | 33.7 (30.6 to 37.5) | 39.9 (36.2 to 44.7) |
| Flavanones to mg/d | 0.5 (0.1 to 1.3) | 0.9 (0.3 to 12.7) | 0.9 (0.3 to 14.1) | 0.8 (0.2 to 12.9) | 1.8 (0.7 to 23.5) |
| Polymers to mg/d | 102.2 (47.6 to 169.0) | 317.8 (246.3 to 386.0) | 513.9 (445.2 to 577.3) | 692.2 (635.5 to 738.1) | 838.1 (757.6 to 951.8) |
| Proanthocyanidins to mg/d<sup>a</sup> | 97.5 (54.5 to 151.5) | 175.7 (122.5 to 252.8) | 210.7 (159.4 to 287.4) | 232.1 (185.6 to 306.6) | 373.0 (289.3 to 518.9) |
| Flavodiet score (tea capped at 4 cups) to servings/d<sup>b</sup> | 1.2 (0.6 to 1.8) | 2.7 (2.2 to 3.2) | 3.8 (3.3 to 4.3) | 4.7 (4.2 to 5.2) | 5.9 (5.2 to 6.8) |
| Tea | 0.0 (0.0 to 0.3) | 1.3 (0.5 to 2.0) | 2.3 (1.8 to 3.0) | 3.5 (2.8 to 4.0) | 4.0 (3.3 to 4.0) |
| Red wine | 0.0 (0.0 to 0.0) | 0.0 (0.0 to 0.7) | 0.0 (0.0 to 0.8) | 0.0 (0.0 to 0.7) | 0.6 (0.0 to 1.5) |
| Apples | 0.0 (0.0 to 0.5) | 0.2 (0.0 to 0.6) | 0.2 (0.0 to 0.7) | 0.2 (0.0 to 0.6) | 0.5 (0.0 to 1.0) |
| Berries | 0.0 (0.0 to 0.1) | 0.0 (0.0 to 0.2) | 0.0 (0.0 to 0.2) | 0.0 (0.0 to 0.2) | 0.0 (0.0 to 0.2) |
| Grapes | 0.0 (0.0 to 0.2) | 0.0 (0.0 to 0.2) | 0.0 (0.0 to 0.2) | 0.0 (0.0 to 0.2) | 0.0 (0.0 to 0.3) |
| Grapefruit | 0.0 (0.0 to 0.0) | 0.0 (0.0 to 0.0) | 0.0 (0.0 to 0.0) | 0.0 (0.0 to 0.0) | 0.0 (0.0 to 0.0) |
| Oranges | 0.0 (0.0 to 0.0) | 0.0 (0.0 to 0.0) | 0.0 (0.0 to 0.0) | 0.0 (0.0 to 0.0) | 0.0 (0.0 to 0.2) |
| Peppers | 0.0 (0.0 to 0.1) | 0.0 (0.0 to 0.1) | 0.0 (0.0 to 0.1) | 0.0 (0.0 to 0.1) | 0.0 (0.0 to 0.1) |
| Onions | 0.0 (0.0 to 0.2) | 0.0 (0.0 to 0.2) | 0.1 (0.0 to 0.2) | 0.0 (0.0 to 0.2) | 0.1 (0.0 to 0.2) |
| Dark chocolate | 0.0 (0.0 to 0.0) | 0.0 (0.0 to 0.0) | 0.0 (0.0 to 0.0) | 0.0 (0.0 to 0.0) | 0.0 (0.0 to 0.0) |
| Energy to kcal/d | 1900.5 (1614.3 to 2227.4) | 1972.4 (1682.6 to 2283.0) | 2006.5 (1728.9 to 2317.0) | 2036.8 (1753.8 to 2357.3) | 2125.2 (1832.0 to 2450.0) |
| Fat to g/d | 67.6 (53.7 to 83.8) | 68.9 (55.1 to 85.3) | 70.4 (56.6 to 86.1) | 71.9 (57.7 to 87.9) | 72.4 (57.8 to 88.8) |
| Alcohol not from red wine to g/d | 5.1 (1.6 to 13.7) | 5.1 (1.7 to 13.3) | 5.1 (1.7 to 13.7) | 5.1 (1.7 to 12.0) | 5.1 (1.7 to 12.0) |
| hPDI score | 59.2 (56.0 to 62.5) | 60.5 (57.3 to 63.4) | 60.8 (57.7 to 63.5) | 61.0 (58.0 to 64.0) | 62.5 (59.5 to 65.2) |
| Dietary assessment completion to d | 3.0 (2.0 to 4.0) | 3.0 (2.0 to 4.0) | 3.0 (2.0 to 4.0) | 3.0 (2.0 to 4.0) | 3.0 (2.0 to 4.0) |
| **Demographic** |  |  |  |  |  |
| Age to y | 58.4 (50.9 to 64.4) | 59.9 (52.5 to 65.2) | 60.5 (53.2 to 65.5) | 61.0 (54.0 to 65.6) | 61.7 (55.1 to 66.0) |
| Duration of follow-up to y | 10.5 (10.4 to 10.9) | 10.5 (10.4 to 10.9) | 10.5 (10.4 to 10.9) | 10.5 (10.4 to 10.9) | 10.5 (10.4 to 10.9) |
| Physical activity to No. (%) |  |  |  |  |  |
| Irregular | 10 259 (44.7) | 9726 (42.3) | 9650 (42.0) | 9786 (42.6) | 9062 (39.5) |
| Regular | 11 905 (51.8) | 12 543 (54.6) | 12 615 (54.9) | 12 461 (54.3) | 13 253 (57.7) |
| Unknown | 806 (3.5) | 700 (3.0) | 705 (3.1) | 722 (3.1) | 655 (2.9) |
| BMI to kg/m<sup>2</sup> | 26.4 (23.7 to 29.7) | 25.9 (23.5 to 28.9) | 25.8 (23.5 to 28.7) | 25.8 (23.4 to 28.7) | 25.9 (23.6 to 28.6) |
| Sex to No. (%) |  |  |  |  |  |
| Female | 13 473 (58.7) | 12 777 (55.6) | 12 552 (54.6) | 12 587 (54.8) | 11 904 (51.8) |
| Male | 9497 (41.3) | 10 192 (44.4) | 10 418 (45.4) | 10 382 (45.2) | 11 066 (48.2) |
| Race and ethnicity to No. (%) |  |  |  |  |  |
| Asian | 207 (0.9) | 292 (1.3) | 266 (1.2) | 143 (0.6) | 98 (0.4) |
| Black | 373 (1.6) | 246 (1.1) | 127 (0.6) | 61 (0.3) | 34 (0.1) |
| Chinese | 88 (0.4) | 82 (0.4) | 64 (0.3) | 45 (0.2) | 18 (0.1) |
| White | 21 865 (95.2) | 21 980 (95.7) | 22 198 (96.6) | 22 474 (97.8) | 22 610 (98.4) |
| Multiracial | 175 (0.8) | 120 (0.5) | 129 (0.6) | 88 (0.4) | 66 (0.3) |

**Table 1. Dietary intake and baseline characteristics by quintiles of total flavonoid intake (continued)**
| Characteristics | Value, median (IQR) |  |  |  |  |
| --- | --- | --- | --- | --- | --- |
|  | Quintile 1 (lowest)<br>(n = 22 970) | Quintile 2<br>(n = 22 969) | Quintile 3<br>(n = 22 970) | Quintile 4<br>(n = 22 969) | Quintile 5 (highest)<br>(n = 22 970) |
| Other<sup>c</sup> | 183 (0.8) | 177 (0.8) | 113 (0.5) | 85 (0.4) | 68 (0.3) |
| Unknown | 79 (0.3) | 72 (0.3) | 73 (0.3) | 73 (0.3) | 76 (0.3) |
| Townsend deprivation index,<br>No. (%) |  |  |  |  |  |
| Low | 4103 (17.9) | 4358 (19.0) | 4721 (20.6) | 4790 (20.9) | 4979 (21.7) |
| Moderate | 13 339 (58.1) | 13 662 (59.5) | 13 826 (60.2) | 13 890 (60.5) | 14 113 (61.4) |
| High | 5496 (23.9) | 4924 (21.4) | 4399 (19.2) | 4267 (18.6) | 3856 (16.8) |
| Unknown | 32 (0.1) | 25 (0.1) | 24 (0.1) | 22 (0.1) | 22 (0.1) |
| Education level, No. (%) |  |  |  |  |  |
| Lower secondary | 3308 (14.4) | 3003 (13.1) | 3072 (13.4) | 3238 (14.1) | 2986 (13.0) |
| Upper secondary | 1498 (6.5) | 1405 (6.1) | 1437 (6.3) | 1441 (6.3) | 1353 (5.9) |
| Vocational | 2565 (11.2) | 2316 (10.1) | 2365 (10.3) | 2516 (11.0) | 2491 (10.8) |
| Higher | 14 109 (61.4) | 14 919 (65.0) | 14 560 (63.4) | 13 949 (60.7) | 14 562 (63.4) |
| Other | 1421 (6.2) | 1237 (5.4) | 1478 (6.4) | 1743 (7.6) | 1519 (6.6) |
| Unknown | 69 (0.3) | 89 (0.4) | 58 (0.3) | 82 (0.4) | 59 (0.3) |
| Smoking, No. (%) |  |  |  |  |  |
| Ever | 9498 (41.3) | 9685 (42.2) | 9636 (42.0) | 9623 (41.9) | 10 085 (43.9) |
| Never | 13 420 (58.4) | 13 239 (57.6) | 13 287 (57.8) | 13 296 (57.9) | 12 832 (55.9) |
| Unknown | 52 (0.2) | 45 (0.2) | 47 (0.2) | 50 (0.2) | 53 (0.2) |
| Sleep duration, No. (%) |  |  |  |  |  |
| >8 | 1365 (5.9) | 1341 (5.8) | 1277 (5.6) | 1327 (5.8) | 1314 (5.7) |
| 7-8 | 16 256 (70.8) | 16 769 (73.0) | 16 909 (73.6) | 16 859 (73.4) | 16 993 (74.0) |
| ≤ 6 | 5294 (23.0) | 4811 (20.9) | 4740 (20.6) | 4744 (20.7) | 4610 (20.1) |
| Unknown | 55 (0.2) | 48 (0.2) | 44 (0.2) | 39 (0.2) | 53 (0.2) |
| Family history of depression,<br>No. (%) |  |  |  |  |  |
| Yes | 2973 (12.9) | 2940 (12.8) | 2894 (12.6) | 2986 (13.0) | 2932 (12.8) |
| No | 19 997 (87.1) | 20 029 (87.2) | 20 076 (87.4) | 19 983 (87.0) | 20 038 (87.2) |
| No. of medications taken,<br>No. (%) |  |  |  |  |  |
| 0 | 11 868 (51.7) | 11 948 (52.0) | 11 792 (51.3) | 11 616 (50.6) | 11 541 (50.2) |
| 1 | 7102 (30.9) | 6982 (30.4) | 7043 (30.7) | 7148 (31.1) | 7020 (30.6) |
| 2 | 2417 (10.5) | 2477 (10.8) | 2570 (11.2) | 2559 (11.1) | 2637 (11.5) |
| 3 | 1291 (5.6) | 1281 (5.6) | 1277 (5.6) | 1370 (6.0) | 1457 (6.3) |
| >4 | 292 (1.3) | 281 (1.2) | 288 (1.3) | 276 (1.2) | 315 (1.4) |
| No. of chronic conditions,<br>No. (%) |  |  |  |  |  |
| 0 | 21 414 (93.2) | 21 380 (93.1) | 21 297 (92.7) | 21 231 (92.4) | 21 137 (92.0) |
| 1 | 1502 (6.5) | 1542 (6.7) | 1606 (7.0) | 1693 (7.4) | 1776 (7.7) |
| >2 | 54 (0.2) | 47 (0.2) | 67 (0.3) | 45 (0.2) | 57 (0.2) |

Abbreviations: BMI, body mass index; hPDI, healthful plant-based diet index.
<sup>a</sup> Proanthocyanidins are also included in the polymer subclass.
<sup>b</sup> Flavodiet score was calculated by summing intakes (in servings per day) of tea (black and green), red wine, apples, berries, grapes, oranges, grapefruit, sweet peppers, onions, and dark chocolate.
<sup>c</sup> Other race and ethnicity was self-selected and includes no additional information.

### Hierarchical levels of flavonoid intake and depression

During a median follow-up of 10.5 years (IQR, 10.4 to 10.9 years), which accumulated 1 182 800 person-years, 2965 cases of incident depression were observed. As detailed in **Table 2**, both total flavonoid intake and the Flavodiet Score were associated with a reduced risk of depression. After full adjustment, the corresponding HRs (95% CIs) across increasing quintiles of total flavonoid intake were 1.00 (ref), 0.89 (0.80, 1.00), 0.93 (0.83 to 1.04), 0.94 (0.84 to 1.05), and 0.84 (0.75 to 0.95) (P _trend_ < 0.05), with HR per SD (95% CI) being 0.95 (0.91 to 0.98). For the Flavodiet Score, each three servings increase was associated with a 10% lower risk of depression (HR: 0.90; 95% CI: 0.95 to 0.96). Linear associations were further suggested by RCS curves for total flavonoid intake and Flavodiet Score with depression risk (*P* _non-linearity_ > 0.05 for both) (**Supplementary Figure 4**).

**Table 2.**
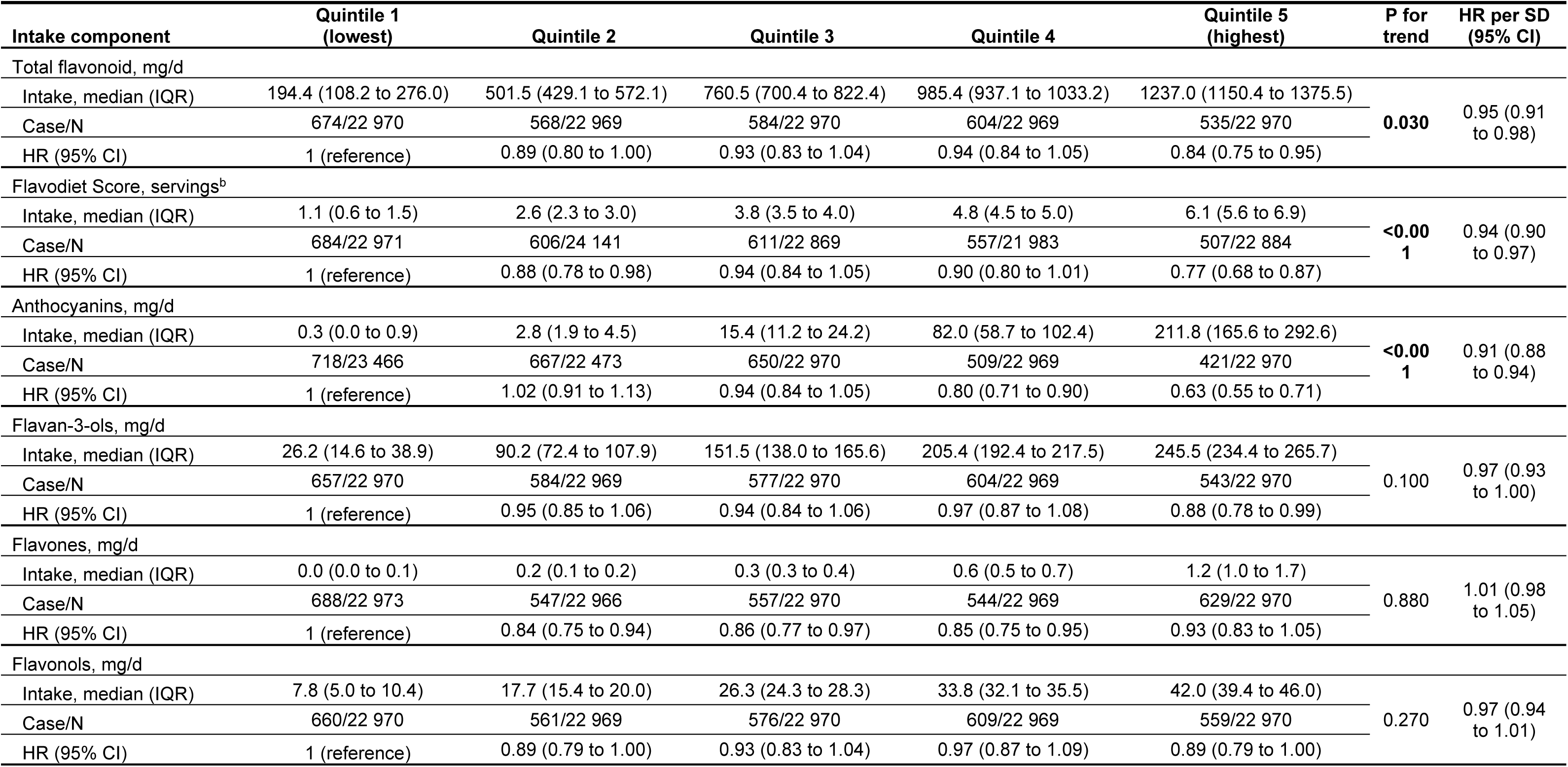
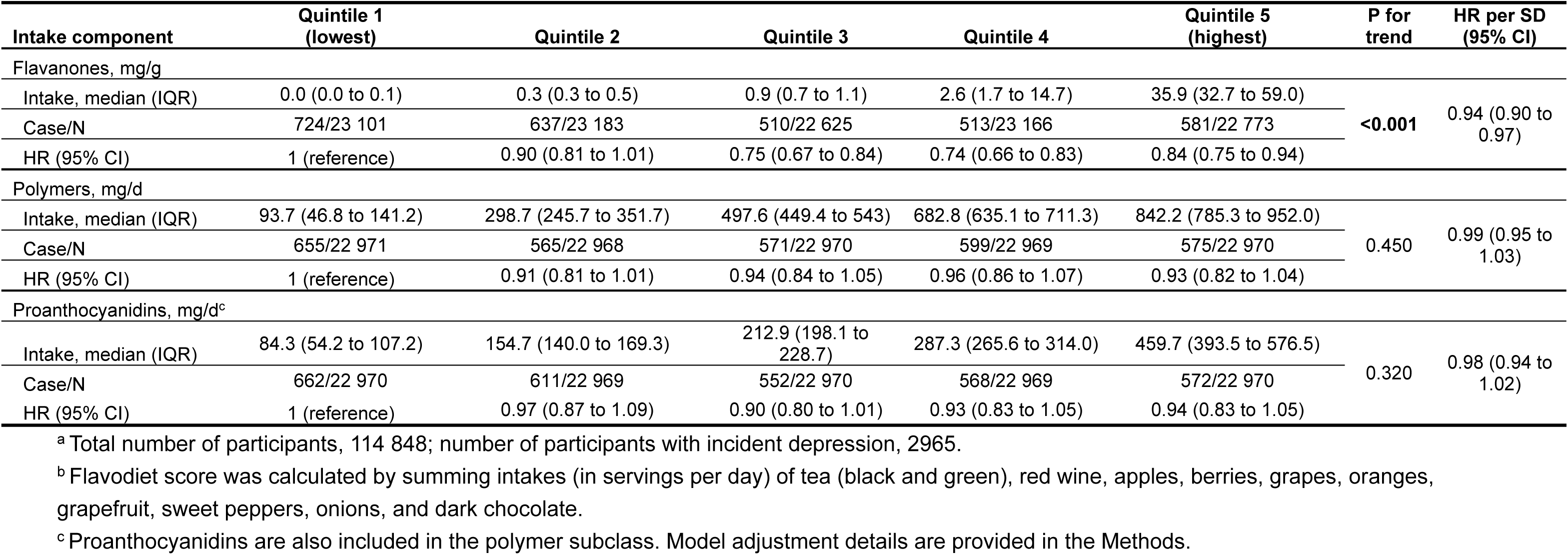
Risk of depression by quintiles of total flavonoid, Flavodiet Score and flavonoid subclass intake^a^.

Among the flavonoid subclasses (**Table 2**), the intakes of anthocyanins and flavanones were linearly associated with a reduced risk of depression (*P* _trend_ < 0.001 for both), with HRs (95% CIs) of higher intakes (quintile 5 vs. 1) being 0.63 (0.55 to 0.71) and 0.84 (0.75 to 0.94), respectively. Furthermore, a decreased risk of depression was observed in certain higher intake groups for both flavan-3-ols and flavones compared with their respective lowest quintiles, although no linear trend was exhibited across the quintiles for either subclass. The Pearson Correlation Coefficients among total flavonoid intake, Flavodiet Score, and intakes of flavonoid subclasses ranged from 0.06 to 0.87 (**Supplementary Figure 5**), and the correlation matrix among flavonoid compounds was presented in **Supplementary Figure 6**.

Regarding specific flavonoid compounds (**Table 3**), intakes corresponding to Cluster 1, Cluster 5, and Cluster 6 were linearly associated with a reduced risk of depression (P _trend_ <0.001 for all), as determined by the k-means method, with corresponding HRs (95% CIs) of higher intakes (quintiles 5 vs. 1) being 0.82 (0.72 to 0.92), 0.63 (0.56 to 0.71), and 0.78 (0.69 to 0.88), respectively. Cluster 1 comprised apigenin, hesperetin, isorhamnetin, and naringenin. Cluster 5 consisted of four anthocyanins (delphinidin, malvidin, peonidin, and petunidin), and Cluster 6 included proanthocyanidin dimers, catechin, and epicatechin (**Supplementary Table 4 & Supplementary Figure 7**). The top two food items with the highest content of each compound within these three clusters were summarized in **Supplementary Table 5**.

**Table 3.** Risk of depression by quintiles of flavonoid intake clusters derived from k-means method^a,b^.

| K-means cluster | Quintile 1 (lowest) | Quintile 2 | Quintile 3 | Quintile 4 | Quintile 5 (highest) | P for trend | HR per SD (95% CI) |
| --- | --- | --- | --- | --- | --- | --- | --- |
| Cluster 1 |  |  |  |  |  |  |  |
| Case/N | 694/22 986 | 666/22 953 | 617/22 970 | 454/22 969 | 534/22 970 | <0.001 | 0.91 (0.88 to 0.95) |
| HR (95% CI) | 1 (reference) | 0.97 (0.87 to 1.09) | 0.94 (0.84 to 1.05) | 0.70 (0.62 to 0.80) | 0.82 (0.72 to 0.92) |  |  |
| Cluster 2 |  |  |  |  |  |  |  |
| Case/N | 675/22 970 | 561/22 969 | 588/22 970 | 541/22 969 | 600/22 970 | 0.210 | 0.94 (0.91 to 0.97) |
| HR (95% CI) | 1 (reference) | 0.88 (0.79 to 0.99) | 0.93 (0.83 to 1.04) | 0.85 (0.76 to 0.96) | 0.98 (0.86 to 1.10) |  |  |
| Cluster 3 |  |  |  |  |  |  |  |
| Case/N | 645/22 970 | 584/22 969 | 574/22 970 | 562/22 969 | 600/22 970 | 0.380 | 1.01 (0.97 to 1.05) |
| HR (95% CI) | 1 (reference) | 0.97 (0.87 to 1.09) | 0.99 (0.88 to 1.11) | 0.97 (0.86 to 1.09) | 1.04 (0.92 to 1.17) |  |  |
| Cluster 4 |  |  |  |  |  |  |  |
| Case/N | 640/22 970 | 570/22 969 | 589/22 970 | 584/22 969 | 582/22 970 | 0.610 | 0.98 (0.95 to 1.02) |
| HR (95% CI) | 1 (reference) | 0.95 (0.85 to 1.06) | 0.99 (0.89 to 1.11) | 0.98 (0.88 to 1.10) | 0.95 (0.84 to 1.06) |  |  |
| Cluster 5 |  |  |  |  |  |  |  |
| Case/N | 740/23 851 | 666/22 567 | 642/22 493 | 493/22 967 | 424/22 970 | <0.001 | 0.86 (0.83 to 0.89) |
| HR (95% CI) | 1 (reference) | 1.02 (0.92 to 1.14) | 0.93 (0.84 to 1.04) | 0.77 (0.69 to 0.87) | 0.63 (0.56 to 0.71) |  |  |
| Cluster 6 |  |  |  |  |  |  |  |
| Case/N | 718/22 970 | 612/22 969 | 597/22 970 | 508/22 969 | 530/22 970 | <0.001 | 0.93 (0.89 to 0.97) |
| HR (95% CI) | 1 (reference) | 0.88 (0.79 to 0.98) | 0.88 (0.79 to 0.98) | 0.76 (0.67 to 0.85) | 0.78 (0.69 to 0.88) |  |  |
| Cluster 7 |  |  |  |  |  |  |  |
| Case/N | 652/22 970 | 574/22 969 | 567/22 974 | 537/22 965 | 635/22 970 | 0.620 | 0.99 (0.95 to 1.02) |
| HR (95% CI) | 1 (reference) | 0.98 (0.88 to 1.10) | 0.94 (0.84 to 1.06) | 0.90 (0.80 to 1.01) | 1.01 (0.90 to 1.14) |  |  |

<sup>a</sup> Total number of participants, 114 848; number of participants with incident depression, 2965.
<sup>b</sup> Model adjustment details are provided in the Methods.

### Sensitivity, stratified, and mediation analysis

The associations between higher intake of total flavonoids, anthocyanins, and flavanones and a lower depression risk were not markedly altered in sensitivity analyses. These analyses included participants aged > 60 years at baseline (**Supplementary Table 6**), those diagnosed with depression within 2 years of follow-up excluded (**Supplementary Table 7**), those with at least 5 years of follow-up (**Supplementary Table 8**), and those residing in areas of high socioeconomic deprivation or low levels of education (**Supplementary Table 9**). Subgroup analyses, stratified by covariates in the fully adjusted models, yielding HRs ranging from 0.91 to 1.08, suggesting no evidence of significant interaction (**Supplementary Figure 8**). The associations between total flavonoid intake and depression were displayed for females and males, respectively in **Supplementary Table 10 & Supplementary Table 11**.

Physical measures, biochemical indices, and chronic disease history that were significantly associated with total flavonoid intake were selected as potential mediators (**Supplementary Table 12**). The top five factors exhibiting the largest absolute mediating effects on the association between total flavonoid intake and depression risk (**Table 4**) were creatinine to cystatin C ratio (Cr/CysC ratio), MQI, hand grip strength, apolipoprotein A, and HDL-cholesterol, with mediation effects ranging from 13.3% to 4.2%. Regarding the history of chronic diseases, a significant mediating effect was found for endocrine system diseases. Further analyses demonstrated that diabetes was the main driven factor, with a mediation proportion of 3.7% (*P* < 0.05). Path diagrams further suggested a sequential chain of intermediate factors, representing the influence of flavonoid intake on depression via Cr/CysC ratio or MQI and subsequent diabetes status (**Figure 1**).

**Figure 1.**
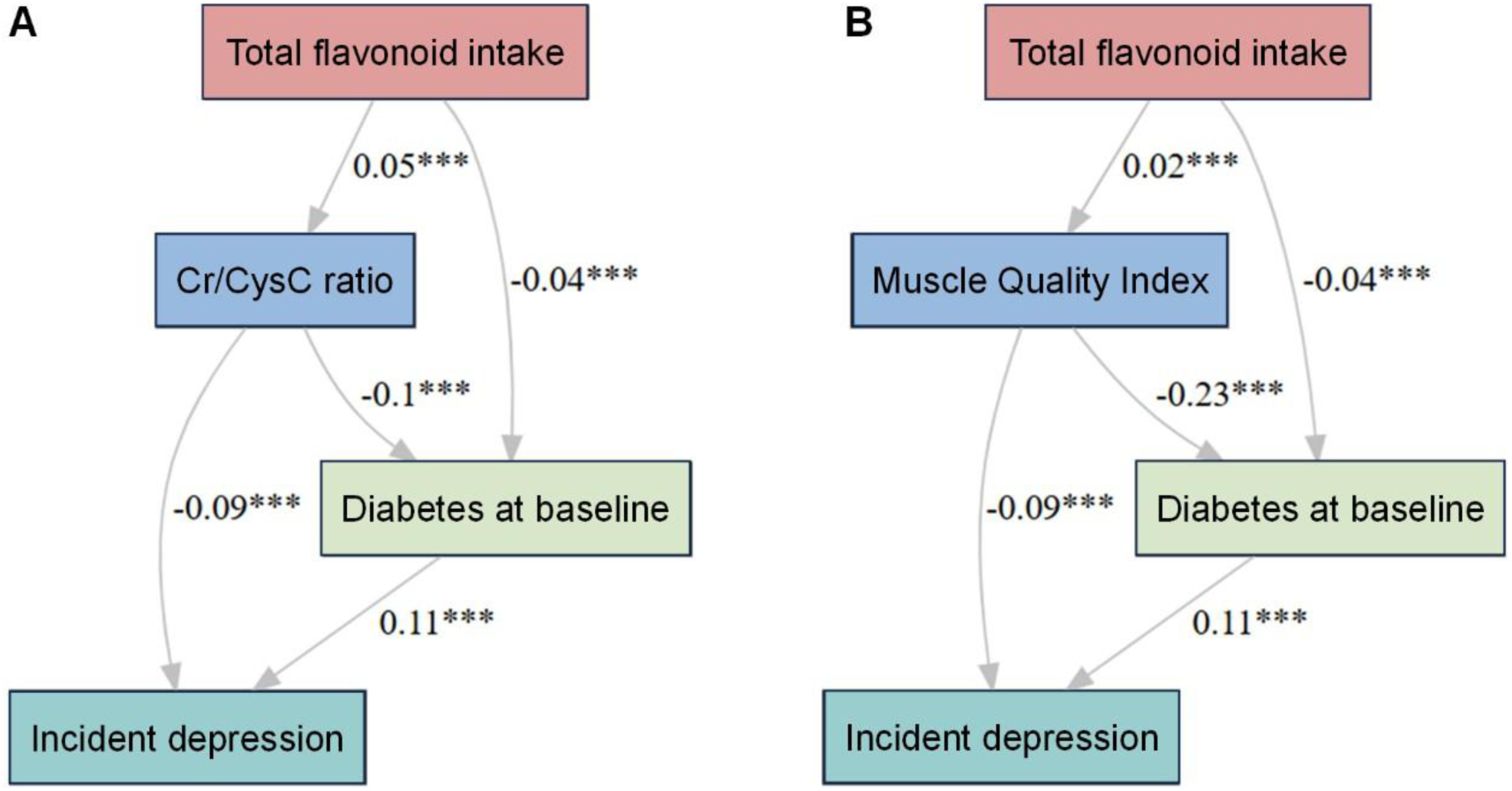
Path diagram showing the observed direct effects between two largest potential mediators and diabetes in the association of total flavonoid with depression using path analysis^a,b,c^. ^a^ Total number of participants, 106 924; number of participants with incident depression, 2730. ^b^ Total number of participants, 112 921; number of participants with incident depression, 2880. ^c^ Muscle quality index, calculated as the ratio of dominant hand grip strength to the corresponding arm fat-free.

**Table 4.** Mediation analysis for the association between total flavonoid intake and depression risk^a^.

|  | Estimate |  |  |  | P value for |
| --- | --- | --- | --- | --- | --- |
|  | Total Effect | ACME | ADE | Mediated Prop. (%) | Mediated Prop. |
| **Biochemical indices** |  |  |  |  |  |
| Alanine aminotransferase | -3.15E-03 | 5.58E-06 | -3.15E-03 | -0.2 | 0.700 |
| Alkaline phosphatase | -3.17E-03 | 5.60E-05 | -3.23E-03 | -1.8 | **0.048** |
| Apolipoprotein A | -3.26E-03 | -1.50E-04 | -3.11E-03 | 4.6 | 0.052 |
| Apolipoprotein B | -3.16E-03 | 5.07E-05 | -3.21E-03 | -1.6 | 0.160 |
| Calcium | -3.32E-03 | 6.90E-06 | -3.32E-03 | -0.2 | 0.848 |
| Creatinine | -3.17E-03 | 1.54E-05 | -3.18E-03 | -0.5 | 0.680 |
| Creatine to cystatin-C ratio | -3.22E-03 | -4.29E-04 | -2.39E-03 | 13.3 | **0.036** |
| Cystatin C | -3.23E-03 | -9.40E-05 | -3.13E-03 | 2.9 | **0.040** |
| Direct bilirubin | -2.65E-03 | -7.38E-06 | -2.64E-03 | 0.3 | 0.744 |
| Gamma glutamyltransferase | -3.22E-03 | 5.51E-05 | -3.28E-03 | -1.7 | 0.060 |
| Glucose | -3.31E-03 | 1.74E-05 | -3.33E-03 | -0.5 | 0.300 |
| Glycated haemoglobin | -2.51E-03 | -3.38E-05 | -2.47E-03 | 1.3 | 0.064 |
| HDL-cholesterol | -3.27E-03 | -1.36E-04 | -3.13E-03 | 4.2 | 0.104 |
| IGF-1 | -3.29E-03 | -6.99E-05 | -3.22E-03 | 2.1 | **0.020** |
| LDL direct | -3.06E-03 | 4.71E-05 | -3.11E-03 | -1.5 | 0.072 |
| Phosphate | -3.21E-03 | 8.02E-05 | -3.29E-03 | -2.5 | 0.460 |
| SHBG | -3.23E-03 | -3.32E-05 | -3.19E-03 | 1.0 | 0.052 |
| Total bilirubin | -3.07E-03 | -3.48E-05 | -3.03E-03 | 1.1 | 0.084 |
| Total protein | -3.26E-03 | -3.87E-05 | -3.22E-03 | 1.2 | 0.100 |
| Triglycerides | -3.07E-03 | -4.50E-05 | -3.02E-03 | 1.5 | 0.364 |
| Urate | -3.22E-03 | -1.12E-05 | -3.21E-03 | 0.3 | 0.364 |
| Urea | -3.17E-03 | -6.41E-05 | -3.11E-03 | 2.0 | 0.160 |
| Vitamin D | -3.24E-03 | -4.34E-07 | -3.24E-03 | <0.1 | 0.976 |
| **Physical measures** |  |  |  |  |  |
| Arm fat free mass (left) | -3.28E-03 | -8.85E-06 | -3.27E-03 | 0.3 | 0.580 |
| Arm fat free mass (right) | -3.30E-03 | 4.96E-07 | -3.30E-03 | >-0.1 | 0.980 |
| Arm fat mass (left) | -3.16E-03 | -1.23E-04 | -3.03E-03 | 3.9 | **0.040** |
| Arm fat mass (right) | -3.18E-03 | -1.23E-04 | -3.05E-03 | 3.9 | **0.036** |
| Arm fat percentage (left) | -3.21E-03 | -7.62E-05 | -3.14E-03 | 2.4 | **0.040** |
| Arm fat percentage (right) | -3.21E-03 | -5.68E-05 | -3.16E-03 | 1.8 | 0.052 |
| Arm predicted mass (left) | -3.28E-03 | -8.43E-06 | -3.27E-03 | 0.3 | 0.540 |
| Arm predicted mass (right) | -3.30E-03 | 3.62E-06 | -3.31E-03 | -0.1 | 0.832 |
| Basal metabolic rate | -3.29E-03 | -2.81E-05 | -3.26E-03 | 0.9 | 0.268 |
| Hand grip strength | -3.55E-03 | -1.89E-04 | -3.36E-03 | 5.3 | **0.016** |
| Leg fat free mass (left) | -3.29E-03 | -2.23E-05 | -3.27E-03 | 0.7 | 0.460 |
| Leg fat free mass (right) | -3.29E-03 | -1.77E-05 | -3.27E-03 | 0.5 | 0.496 |
| Leg fat mass (left) | -3.16E-03 | -1.07E-04 | -3.05E-03 | 3.4 | **0.036** |
| Leg fat mass (right) | -3.16E-03 | -1.08E-04 | -3.05E-03 | 3.4 | **0.044** |
| Leg predicted mass (left) | -3.29E-03 | -2.40E-05 | -3.27E-03 | 0.7 | 0.356 |
| Leg predicted mass (right) | -3.29E-03 | -1.65E-05 | -3.27E-03 | 0.5 | 0.472 |
| Muscle Mass Index | -3.35E-03 | -2.17E-04 | -3.13E-03 | 6.5 | **0.020** |
| Trunk fat free mass | -3.30E-03 | -5.44E-06 | -3.29E-03 | 0.2 | 0.728 |

**Table 4. Mediation analysis for the association between total flavonoid intake and depression risk<sup>a</sup> (continued)**
|  | Estimate |  |  |  | P value for Mediated Prop. |
| --- | --- | --- | --- | --- | --- |
|  | Total Effect | ACME | ADE | Mediated Prop. (%) |  |
| Trunk fat mass | -3.22E-03 | -6.08E-05 | -3.15E-03 | 1.9 | **0.040** |
| Trunk predicted mass | -3.30E-03 | -5.68E-06 | -3.29E-03 | 0.2 | 0.628 |
| Whole body fat free mass | -3.31E-03 | -1.12E-05 | -3.29E-03 | 0.3 | 0.592 |
| Whole body fat mass | -3.23E-03 | -1.20E-04 | -3.13E-03 | 3.2 | **0.028** |
| Whole body water mass | -3.30E-03 | -9.61E-06 | -3.29E-03 | 0.3 | 0.568 |
| **History of chronic diseases** |  |  |  |  |  |
| Diseases of endocrine system | -1.22E-03 | -3.31E-05 | -1.19E-03 | 2.6 | **0.008** |

<sup>a</sup> Model adjustment details are provided in the Methods.

### Flavonoid intake and brain structural change

For the association of total flavonoid intake and the Flavodiet Score with brain structural changes over a median 27.4 months of interval, a secondary analysis was conducted using data from participants with two brain MRI scans. A comparison of their characteristics against those without repeated MRI scans was presented in **Supplementary Table 13**, showing similar levels of total, the Flavodiet Score, and all subclasses of flavonoid, except for flavanones. Brain regions exhibiting structural changes significantly associated with total flavonoid intake (per SD) or Flavodiet Score (per serving) were annotated in **Figure 2**, with corresponding brain volumes at Phase 1 adjusted. Furthermore, brain region changes demonstrating a significant linear trend across the quintiles of total flavonoid intake included less atrophy in the left Caudal anterior cingulate, left Supramarginal, right Caudal middle frontal, and right Precentral, and a smaller increase in MD in the left Anterior Thalamic Radiation tract (**Supplementary Table 14**). The corresponding brain region changes observed for Flavodiet Score included less atrophy in the left Caudal anterior cingulate, left Inferior temporal, left Supramarginal, and right Caudal middle frontal regions (**Supplementary Table 15**).

**Figure 2.**
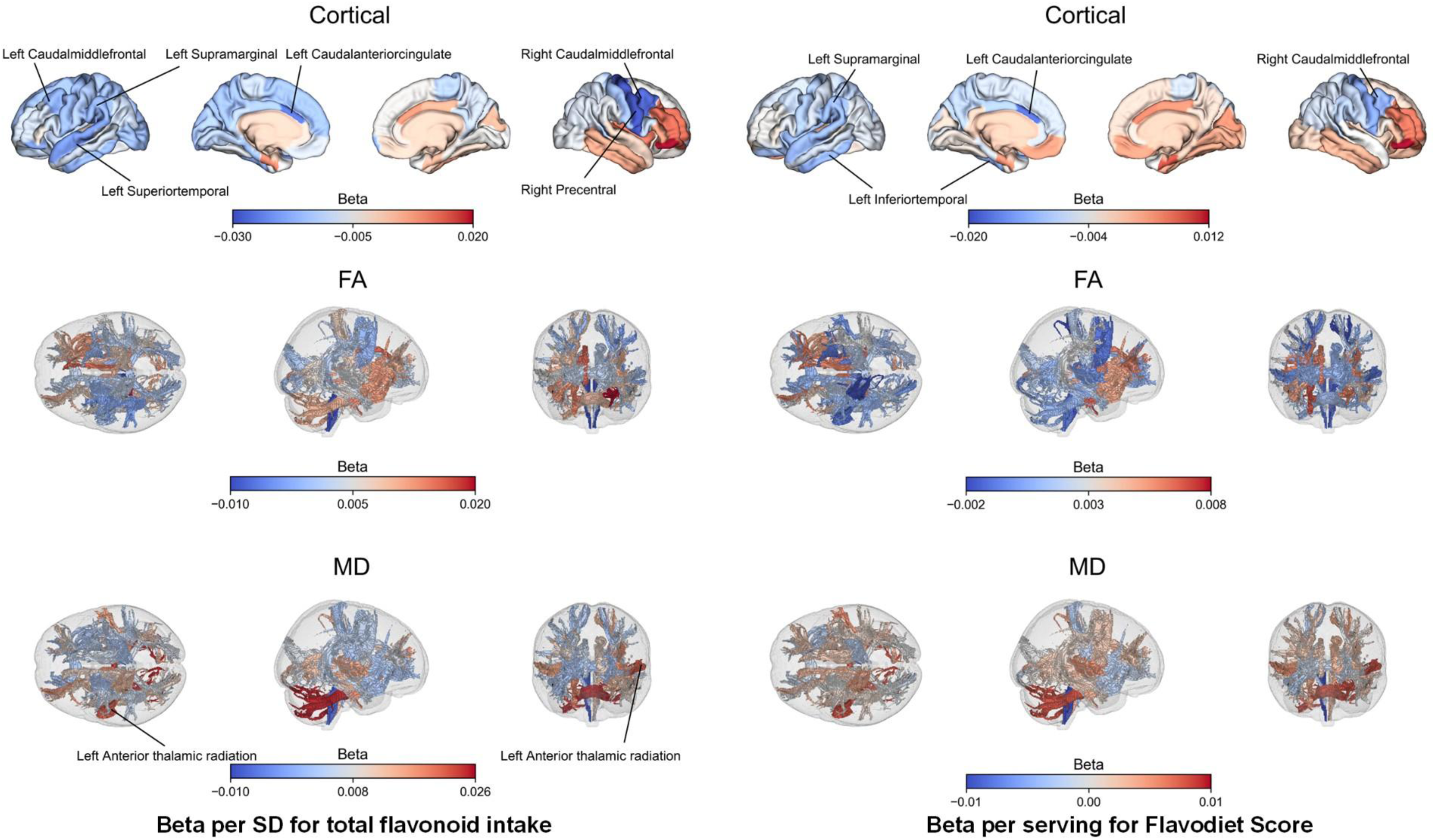
Associations of flavonoid levels with brain structural changes^a,b^. ^a^ Blue denotes decreased volume and red denotes increased volume. ^b^ Total number of participants, 2120. Model adjustment details are provided in the Methods.

## Discussion

The findings of the present study demonstrated that higher flavonoid intake was linearly associated with a reduced risk of depression, whether measured as total flavonoid intake or the Flavodiet Score. This association was particularly driven by anthocyanins and flavanones among flavonoid subclasses. Regarding specific flavonoid compounds, the k-means clustering analysis suggested a linear association with a lower depression risk for Cluster 1 (predominantly driven by flavanones), Cluster 5 (four of six anthocyanins), and Cluster 6 (proanthocyanidin dimers, catechin, and epicatechin). No significant interaction was observed for the association of total flavonoid intake with depression risk by potential effect modifiers, such as demographic, lifestyle, and chronic disease factors.

Furthermore, several biochemical indices, physical measures, and a history of diabetes were found to significantly mediate the association between total flavonoid intake and depression risk, with the Cr/CysC ratio being the most prominent mediator. In terms of brain structure, higher total flavonoid intake was associated with beneficial depression-related brain structural changes, specifically less atrophy in the left Caudal anterior cingulate, left Supramarginal, right Caudal middle frontal, and right Precentral, and a smaller increase in MD within the left Anterior Thalamic Radiation tract.

### Comparison with other studies

Our finding that total flavonoid intake was statistically significantly associated with a reduced risk of depression is largely consistent with published literature. According to the findings from 45 985 females in the Nurses’ Health Study (NHS), the HR (95% CI) of incident depression for quintile 5 vs. quintile 1 was 0.88 (0.80 to 0.97) during a 10-year follow-up, an effect magnitude comparable to our find of 0.84 ^[10]^. Evidence from cross-sectional studies analyses of the National Health and Nutrition Examination Survey also supported this association. The odds ratio (OR) (95% CI) of prevalent depression for quartile 4 vs. quartile 1 was reported as 0.75 (0.56 to 1.00) ^[9]^, and the effect size of prevalent perimenopausal depression for quartile 3 vs. quartile 1 was 0.67 (0.46 to 0.99) ^[11]^. Furthermore, NHS findings indicated that quintile 5 of total flavonoid intake compared with quintile 1 was associated with an 11% lower risk of poor mental health, which was defined as a composite outcome using the MF-5 in the SF-36 and Center for Epidemiologic studies Depression 10 score ^[19]^.

Prior research on the association between flavonoid-rich foods and depression risk is generally scarce. Based on results among females from the NHS and NHSⅡ cohorts ^[10]^, citrus fruit and juice combined was the only flavonoid-rich food (among citrus fruits, citrus juice, tea, and onions) significantly associated with a decreased risk of incident depression. A HR (95% CI) of 0.82 (0.74 to 0.91) was reported for consuming ≥ 2 servings/week compared with <1 serving/wk. Furthermore, decreased odds of major depressive disorder (certified in 2014-2015) were observed in quintile 5 of flavonoid-risk fruit consumption (obtained in 1995 and 2000) compared with quintile 1 in the JPHC Saku Mental Health Study (OR: 0.44; 95% CI: 0.20 to 0.97) ^[20]^. The Flavodiet Score, calculated as the sum of main dietary contributors to flavonoid intake, was previously found to be associated with 12% and 18% lower risks of poor mental health when comparing quintile 5 with quintile 1 in US females and males, respectively ^[19]^. The current research displayed a 23% (13%, 32%) lower risk of incident depression when comparing quintile 5 of the Flavodiet Score with quintile 1, in consistent with previous studies.

Our study showed that the associations of total flavonoid intake and the Flavodiet Score with the risk of incident depression were both linear, as supported by both the Chi-square test for trend and the test for non-linearity in RCS. Consistent with our findings, linear trends across quintiles were previously observed for the association of total flavonoid intake with incident depression risk ^[10]^, and the association of total flavonoid intake and the Flavodiet Score with the risk of poor mental health ^[19]^ among females in the NHS cohort. Conversely, no evidence of linear trends across quintiles was suggested for either total flavonoid intake or the Flavodiet Score with the risk of poor mental health among males in the Health Professionals Follow-up Study (HPFS) ^[19]^. Although no sex interaction was found in the current study for the association of total flavonoid intake with incident depression risk, a linear trend was similarly observed in females but not in males. Furthermore, no linear trends were suggested by cross-sectional studies from the NHANES for the association of total flavonoid intake with the risk of depression ^[21]^ or the risk of perimenopausal depression ^[11]^. Although not the sole explanation, these observed discrepancies may be partly due to the reverse causation, where individuals with a higher depression risk might turn to adopt diets with a higher flavonoid intake.

As previously reported in cross-sectional studies from the NHANES, a 33% lower risk ^[22]^ and a 31% lower risk of depression for tertile 3 of anthocyanidins vs. tertile 1 ^[21]^, along with linear trends across tertiles. Consistent with these results, the current prospective study found that anthocyanin intake was linearly associated with a 37% lower risk of incident depression (quintile 5 vs. quintile 1) during a median follow-up of 10.5 years. Evidence from the NHS and NHSⅡ females cohorts showed that quintile 5 of flavanone intake was associated with a 10% lower risk of incident depression, with a linear trend across quintiles during a 10-year follow-up ^[10]^. This observed linear trend for flavanones was consistent with current finding of a 16% lower risk of depression for quintile 5 vs. quintile 1 and it is further supported by cross-sectional NHANES studies ^[9, 11]^. The previously suggested linear association between flavone intake and depression risk ^[10, 9, 21]^, as well as the observed reduced depression risk in specific intake categories of flavan-3-ols ^[10, 9]^, were partially support by the present findings, despite the absence of evidence for linear trends across intake quintiles.

Through a k-means-based clustering approach, our study delineated various flavonoid compounds associated with a lower depression risk, supported by prior literature on these compounds per se mainly from animal and *in silico* studies, or their food sources mainly from population-based research. For the flavanones in Cluster 1, animal studies showed that hesperidin could attenuate chronic stress-induced depression ^[23]^, and naringenin could significantly ameliorate depression-like behavioral alterations induced by olfactory bulbectomy ^[24]^. Furthermore, a cross-sectional analysis from the Mediterranean Healthy Eating, Lifestyle and Aging (MEAL) Study suggested that quartile 4 of naringenin intake vs. quartile 1 was associated with a 51% lower risk of depression ^[25]^. Regarding the anthocyanins from Cluster 5, malvidin-3-O-glucoside was shown to mitigate chronic unpredictable stress-induced depressive behaviors ^[26]^. Additionally, peonidin demonstrated a stronger binding affinity with multiple serotonin receptors than the natural neurotransmitter 5-HT, as evidenced by an *in silico* study ^[27]^. The reason that Cluster 6 containing partial monomers (i.e., catechin and epicatechin) and dimers of proanthocyanidins, instead of Cluster 7 including polymers of proanthocyanidins, was associated with lower depression risk may be partially attributed to decreasing bioavailability. Specifically, a series of *in vivo* and *in vitro* studies confirmed that monomeric and dimeric proanthocyanidins from grapeseed extract could be partially absorbed in the gastrointestinal tract, while orally ingested oligomeric and polymeric proanthocyanidins were not bioavailable prior to interaction with the colonic microbiota ^[28]^. Population-based randomized controlled trials have provided clinical evidence: the intake of orange juice, which is rich in flavanones as hesperidin and narirutin, improved depressive symptoms in young adults with major depressive disorder over eight weeks ^[29]^. Similarly, adolescents supplemented daily with wild blueberry for four weeks reported fewer depression symptoms compared with the placebo group ^[30]^.

Preclinical studies suggests that flavonoids may mitigate depressive behaviors in animal models through various mechanisms. Regarding depression-related molecular targets, quercetin was found to alleviate depression-like behavior by interfering with the binding between α2δ-1 and NMAR1 **^[31]^**. Furthermore, a significant increase in acetylcholinesterase (AchE) and monoamine oxidase (MAO) activity was observed in the cortex and hippocampus following the administration of nanoparticles loaded with hesperidin and quercetin ^[32]^. The antidepressive action of epigallocatechin-3-gallate in mice also involved the upregulation of hippocampal mRNA level of BDNF, the decrease of which is regarded as biomarker of depression pathophysiology ^[33]^. For depression-related neurotransmitters, elevated levels of serotonin, dopamine, and norepinephrine were observed following kaempferol administration in 4T1 mouse BC cells ^[34]^. Prophylactic application of cyanidin was also found to alleviate lipopolysaccharide-induced depression-like behaviors, partially through the attenuation of glutamate-induced excitotoxicity ^[35]^. In terms of central nervous system cells, both quercetin ^[36]^ and baicalin ^[37]^ were found to promote hippocampal neurogenesis via the Wnt/β-catenin pathway. Moreover, quercetin was discovered to inhibit hippocampal microglial activation ^[38]^ and its phenotype transformation, thus protecting white matter from demyelination ^[39]^. Additional mechanisms underlying the effect of flavonoid compounds on depression involve the inhibition of pyroptosis ^[40]^, apoptosis ^[41]^, ferroptosis ^[42]^, neuroinflammation, and oxidative damage ^[43]^, as well as modulation via the microbiota-gut-brain axis ^[44]^.

The ratio of serum Cr/CysC ratio was initially proposed as an indicator of muscle mass by Kashani in 2017 ^[45]^, while the ratio of muscle strength or muscle power per unit of muscle mass was first introduced as a potential marker for muscle quality by Barbat-Artigas in 2012 ^[46]^. Both ratios are established biomarkers for sarcopenia ^[47, 48]^. Prior research from the China Health and Retirement Longitudinal Study (CHARLS) demonstrated that an increase in the Cr/CysC ratio was significantly associated with a decreased risk of depression (OR: 0.69; 95% CI: 0.59 to 0.81 for quartile 4 vs. quartile 1) ^[49]^. Similarly, the UK Biobank study reported that compared with participants in quintile 1 of muscle quality index (MQI), the HR (95% CI) of depression for those in quintile 5 was 0.53 (0.50 to 0.57) ^[48]^. Given the abundant evidence that specific flavonoid compounds, such as epigallocatechin-3-gallate ^[50]^, quercetin ^[51, 52]^, and delphinidin ^[53]^, could reduce muscle atrophy in mouse models by improving mitochondrial homeostatic balance ^[52]^ and alleviating inflammation ^[50]^, it was hypothesized that flavonoids may decrease depression risk through preserving muscle health. This hypothesis was supported by the present results, which showed that 13.3% and 6.5% of the association between total flavonoid intake and depression risk were mediated by the Cr/CysC ratio and MQI, respectively. This is consistent with previous evidence for the mediation role of muscle strength index in the association between adherence to the MedDiet and depression symptoms reported in the Nuts4Brain-Z study ^[54]^. Furthermore, the UKB study has demonstrated that total flavonoid intake was associated with a significantly decreased risk of depression (HR: 0.75; 95% CI: 0.66 to 0.84 for quartile 4 vs. quartile 1) ^[55]^. Moreover, diabetes was associated with a remarkably higher risk of depression (OR: 2.0; 95% CI: 1.8 to 2.2) in a meta-analysis ^[56]^. Our study showed a 3.7% of mediating effect for diabetes in the association of total flavonoid intake and depression risk, which may be through increasing insulin sensitivity ^[57, 58]^ and reshaping the gut microbiota and gut-liver axis ^[59]^ suggested by previous animal research. Our study observed the influence of flavonoid intake via sarcopenia biomarkers and subsequent diabetes status, suggesting a potential role for muscle insulin sensitivity which warrants further confirmation.

Previous investigations into the effects of flavonoid levels on brain structure have been exclusively based on animal models, primarily focusing on reductions in total brain atrophy. For example, cyanidin-3-glucoside, as a flavonoid-based glucoside, was shown to alleviate brain atrophy in naturally aging mice ^[60]^. Only one study, which utilized a tauopathy model based on PS19 mice, examined neocortical volume and demonstrated that a well-characterized senolytic therapy combining dasatinib and quercetin exhibited significant brain atrophy, particularly in the hippocampus and neocortex ^[61]^. In the present study conducted among 2120 participants, no significant association was found between total flavonoid level and reductions in total brain, grey matter, or white matter volume. However, associations between a higher total flavonoid level or a higher Flavodiet Score and less atrophy were observed in specific grey matter regions, including the anterior cingulate, supramarginal, middle frontal, precentral, and inferior temporal cortex, all of which are known to be relevant to depression risk ^[62–65]^. Furthermore, total flavonoid intake was found to be associated with a smaller elevation in MD in the left anterior thalamic radiation tract, suggesting a protective role in white matter integrity. Although all associations between flavonoid intake and brain structural changes became null after multiple testing correction, this work represents the first population-based investigation to explore the relationship between flavonoid levels and both whole-brain and regional grey matter changes. Consequently, these findings provide a crucial foundation for future studies requiring larger sample sizes and longer brain MRI follow-up intervals.

### Strengths and limitations of this study

A major strength of this study lies in the hierarchical examination of flavonoid intake, ranging from total flavonoid and the Flavodiet Score (as the sum of flavonoids-rich food servings) to specific flavonoid subclasses and individual compounds. Furthermore, the mediating roles of biochemical indices, physical measures, and disease history were explored in detail. Finally, the effect of flavonoid intake on brain structural changes was examined among participants with two brain MRI scans. However, several limitations should be acknowledged. Firstly, despite the accumulated average of dietary intake (median 3.0 assessments during a median of 14.0 months period) being used to reduce within-subject variation, known measurement errors and reporting biases inherent to 24-h dietary assessment persist ^[66]^. Secondly, although careful adjustments were made for multiple demographics characteristics, lifestyle factors, clinical conditions, and dietary covariates that may confound the associations of interest, residual confounding remains inevitable given the observational design of this study. Thirdly, most of our study population were Whites and this may limit the generalizability of our findings to other populations in difference race and ethnicities. Fourthly, 24-h dietary recalls are subject to measurement errors due to the self-report nature. Nonetheless, this error is likely non-differential due to the prospective study design, suggesting that the associations may be biased toward the null. Lastly, reverse causation could be a concern, as flavonoid intake may be modified by caregiver intervention shortly before a depression diagnosis. Nevertheless, the associations remained similar when depression cases occurring within 2 years of follow-up were excluded.

### Conclusion and policy implications

A hierarchical analysis of flavonoid intake, including total flavonoids, the Flavodiet Score (servings of flavonoid-rich foods), and specific subclasses (anthocyanins and flavanones) and compounds (suggested by the k-means method), may be linearly associated with a reduced risk of depression. Muscle loss and glucose metabolism dysfunction may play key mediating roles in these associations. Furthermore, increased total flavonoid intake was associated with smaller atrophy in depression-related cortical regions and a reduced increase in the loss of white matter integrity. Supplementation trials involving specific compounds identified in this analysis are warranted by *in vivo* and *in vitro* experiments to clarify the underlying mechanisms. Additionally, randomized controlled clinical trials are required to test the effectiveness of this intervention for lowering depression risk.

## Data Availability

Data from UK Biobank are available on application at www.ukbiobank.ac.uk/register-apply.

## Acknowledgements

This research has been conducted using the UKB Resource (application number 124655). We thank the participants and staff of the UK Biobank for their valuable contributions.

## Contributors

YC, CY, and WL designed the study. WL acquired the data. CY and WL obtained fundings. YC conducted the data analyses and prepared the first draft of the manuscript. YC and QG established the key calculation method of flavonoid intake. HC helped with the statistical method. TL, ZH, AC, EI provided support for other methods. QG, WY, and YL helped with data visualization presentation. All authors contributed to the interpretation of the results and critical revision of the manuscript for important intellectual content and approved the final version of the manuscript. YC, CY, and WL are the guarantors. The corresponding author attests that all listed authors meet authorship criteria and that no others meeting the criteria have been omitted.

## Ethical approval

Ethical approval for the study was granted by the North West Multi-center Research Ethics Committee (REC reference: 21/NW/0157). Informed consent was obtained from all participants via electronic signature at baseline.

