## Supplementary for "Association of Dietary Flavonoid Intake with Incident Depression Risk and Brain Structural Changes: A Prospective Study in the UK Biobank"

### On-line Supplementary Material

**Material list**

**Supplementary Figure 1. Flowchart of the study**

**Supplementary Method 1. Calculation of flavonoid intake**

**Supplementary Table 1. Diagnoses of depression and baseline chronic health conditions**

**Supplementary Method 2. Definition of covariates**

**Supplementary Table 2. The numbers (percentages) of participants with missing covariates**

**Supplementary Figure 2. Histogram of intakes of total flavonoid and subclasses**

**Supplementary Figure 3. Histogram of intakes of flavonoid compounds**

**Supplementary Table 3. Baseline characteristics and dietary intakes by incident depression diagnosis**

**Supplementary Figure 4. Restricted cubic spline for the association of flavonoid levels with depression risk**

**Supplementary Figure 5. Correlation heatmap for intakes of flavonoid subclasses**

**Supplementary Figure 6. Clustered correlation heatmap for intakes of flavonoid compounds**

**Supplementary Table 4. Flavonoid clusters derived from the k-means method**

**Supplementary Figure 7. K-means clustering of flavonoid compound intakes**

**Supplementary Table 5. The top two food items with the highest content of each compound within Cluster1, Cluster 5, and Cluster 6 from the k-means methods**

**Supplementary Table 6. Risk of depression by quintiles of flavonoid intake levels in participants aged more than 60 years**

**Supplementary Table 7. Risk of depression by quintiles of flavonoid intake levels in participants excluding cases diagnosed within 2 years of follow-up**

**Supplementary Table 8. Risk of depression by quintiles of flavonoid intake levels in participants with at least 5 years follow-up**

**Supplementary Table 9. Risk of depression by quintiles of flavonoid intake levels in participants living in areas of high deprivation or with low levels of education**

**Supplementary Figure 8. Subgroup analysis for the association of total flavonoid intake with depression risk**

**Supplementary Table 10. Risk of depression by quintiles of total flavonoid, Flavodiet Score and flavonoid subclass intake among females**

**Supplementary Table 11. Risk of depression by quintiles of total flavonoid, Flavodiet Score and flavonoid subclass intake among males**

**Supplementary Table 12. Association of total flavonoid intake with potential mediators as physical measures, biochemical indices, and history of chronic diseases**

**Supplementary Table 13. Participant characteristics according to the availability of two brain MRI scans**

**Supplementary Table 14. Associations of total flavonoid intake (per SD) with brain structural changes**

**Supplementary Table 15. Associations of the Flavodiet Score (per serving) with brain structural changes**

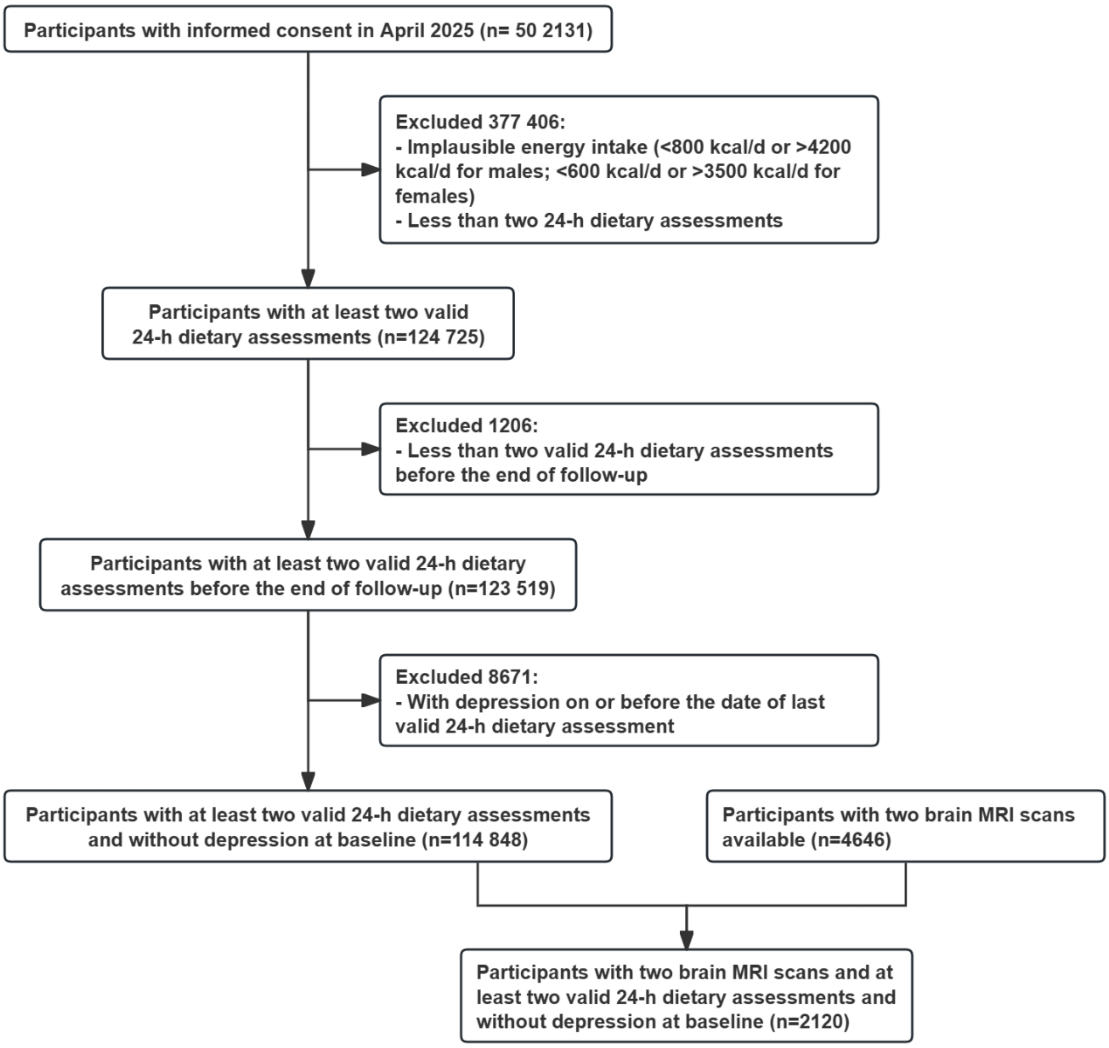


**Supplementary Figure 1. Flowchart of the study**

### Supplementary Method 1. Calculation of flavonoid intake

The ten foods with the highest flavonoid content in the Oxford WebQ were selected based on their contribution to the total intake of flavonoid subclasses in the UKB[1]. These foods were used to calculate the intake of each subclass and compound. The ten selected foods included tea (black and green), apples, berries, red wine, grapes, sweet peppers, onions, dark chocolate, oranges (including satsumas), and grapefruit [2]. The levels of each compound, including flavonoids, proanthocyanidins, and isoflavones, were primarily sourced from the United States Department of Agriculture (USDA) database [3-5]. Data for compounds not available in the USDA database were supplemented using the Phenol-Explorer database [6]. Food items that could not be matched to a flavonoid code in either database were assigned a flavonoid value of zero. The calculation method for the intake of each food item is as follows:

1) **Tea:** Data from two UKB 24-hour dietary recall IDs (100400 and 100420) were used to assess the intake of various tea types [7]. Two NDB numbers (14355 and 99068) were finalized to estimate the content of various compounds based on British tea consumption habits [3]. For green tea, proanthocyanidin and isoflavone content was estimated using NDB numbers 14278 and 99017 [3, 4]. Data pertaining to decaffeinated tea leaves (100470) were categorized based on participants' green tea and black tea consumption [7]. Subsequently, using NDB numbers 14352 (black tea decaffeinated) and 99069 (green tea decaffeinated), the content of each compound was calculated for these categories[3, 4].
2) **Apples:** Apples are a typical fruit rich in flavonoids, with the high concentration of flavonoids in the apple skin making it essential to include them in compound calculations. Referring to previous studies, the proportion of apple peels consumed by European populations was determined to obtain more accurate estimates of compound intake [8]. NDB numbers 09003 and 09004 were used to calculate the compound content of apples with and without peels, respectively. UKB field ID 104450 was used to measure apple intake [3, 4].
3) **Berries:** Berries are a diverse group of fruits that include a wide variety of species, and their definition can vary across academic fields. In this study, the term "soft fruit" was used in its narrow sense to refer specifically to berries. This definition aligns with common dietary patterns and everyday usage, and it provides a clearer categorization within the USDA database [9]. According to the Department for Environment, Food & Rural Affairs (DEFRA) of the UK, different types of berries, including NDB numbers 09302, 09316, 09050, 99653, and 97085, were accounted for based on their consumption in the UK [10, 3, 4]. UKB field ID 104470 was used to measure berry intake. Since NDB number 99653 is missing from the proanthocyanidin database, weights excluding this number were used to calculate proanthocyanidin content [4].
4) **Red wine:** The compound content of red wine is closely linked to the grape varieties used in its production. The proportion of different grape types in the winemaking process is a key factor in determining the specific content of each compound. To estimate the compound content of wines, three NDB numbers (14098, 14097, and

14100) were selected based on Bruwer et al.'s statistics on the grape varieties used for wines in the UK market [3, 4, 11]. UKB field ID 100590 was utilized to assess wine intake.

5) **Grapes:** There are many different types of grapes, and the USDA provides specific compound levels for different grape colors. The International Cool Climate Wine Symposium WineGB Yield Survey was used to match the yields of various grape varieties [12]. Specific compound levels were calculated based on the color information for each variety (NDB numbers 99048, 97074, 99047) [3, 4]. UKB field ID 104500 was used to measure grape intake. Since NDB number 99048 is missing from the proanthocyanidin database, weights excluding this number were used to calculate proanthocyanidin content [4].
6) **Sweet peppers:** Sweet peppers are a well-known dietary source of flavonoids, with different sweet pepper colors containing varying levels of flavonoids. A study on consumer willingness to pay for sweet pepper colors in a European population indicated that color influences purchasing decisions [13]. Based on consumer preferences, green, yellow, and red sweet peppers (NDB numbers 11333, 11821, 11951) were included in the analysis [3, 4]. The intake of sweet peppers was measured using UKB field ID 104290.
7) **Onions:** There are many types of onions in Europe, with varying edible parts. Studies have shown that for spring onions or scallions, both the tops and bulbs (NDB number 11291) are commonly consumed raw [3, 4, 14, 15]. Additionally, boiled onions are widely used in European cuisine (NDB number 11283) [3, 4]. The total flavonoid content of onions was calculated using both raw and boiled onions. UKB field ID 104260 was used to measure onion intake.
8) **Dark chocolate:** Dark chocolate is recognized as a rich source of flavonoids and was therefore included in the analysis [2]. NDB number 99412 and UKB field ID 102290 were used to calculate the flavonoid compound content in dark chocolate [3, 4]. For dark chocolate, proanthocyanidin content was estimated using NDB number 19905 [4].
9) **Oranges:** UKB field IDs 104530 and 104540 were used to measure the intake of whole oranges (including Satsuma) and the corresponding NDB number was 09200 [3, 4].
10) **Grapefruits:** UKB field ID 104490 was used to measure the intake of whole grapefruit and the corresponding NDB number was 99347 [3, 4].
11) **Other food items:** The flavonoid content of the remaining foods was determined using a similar method to that used for the ten highest flavonoid-containing foods [3-6]. When multiple NDB codes corresponded to a single food group, the code representing the variety most widely consumed in the UK was selected. If a food item in the UKB data comprised multiple food groups, the average flavonoid content of the most commonly consumed foods within those groups, based on UK dietary patterns, was calculated. Food items or their variants without corresponding flavonoid data in either database was assigned a flavonoid content of zero. To address inconsistencies between raw and cooked food data in the UKB and the two databases, retention factors were applied based on the USDA assessment model to estimate the flavonoid

content of cooked foods. Specifically, a retention factor of 0.85 was used for all flavonoid subclasses except anthocyanins, which had a retention factor of 0.5, when converting raw food data to cooked food data [16].

The UKB provides data on either the number of servings or the weight of food consumed daily by participants. When explicit food weights were available in the UKB, these data were used directly to calculate individual flavonoid content. Otherwise, food portion sizes were primarily sourced from the Department of Agriculture (UK)[17, 18]. When Department of Agriculture reference standards were unavailable, physical portion sizes obtained from published research, professional dietitians or suppliers were used[19-21]. If a single food item has small, medium, and large portion sizes, the medium portion was used as the representative portion size to increase generalizability.

**Supplementary Table 1. Diagnoses of depression and baseline chronic health conditions**

| Outcome/health conditions | Source of data | Codes |
|---|---|---|
| Depression | Self-report (instance 0 only) | Field ID: 20002 (1286 depression, 1531 post-natal depression) |
| | PHQ-2 (instance 0 only) | Field ID: 2050 (Over the past two weeks, how often have you felt down, depressed or hopeless?), 2060 (Over the past two weeks, how often have you had little interest or pleasure in doing things?) |
| | Hospital inpatient records | ICD-9: 311; ICD-10: F32, F33 |
| | Death registry | ICD-10: F32, F33 |
| Myocardial infarction | Self-report | Field ID: 20002 (1075 heart attack/myocardial infarction) |
| | Hospital inpatient records | ICD-9: 410, 412; ICD-10: I21-I23, I24.1, I25.2 |
| | Death registry | ICD-10: I21-I23, I24.1, I25.2 |
| Stroke | Self-report | Field ID: 20002 (1086 subarachnoid haemorrhage, 1491 brain haemorrhage, 1583 ischaemic stroke) |
| | Hospital inpatient records | ICD-9: 430, 431, 433, 434; ICD-10: I60, I61, I63, I64 |
| | Death registry | ICD-10: I60, I61, I63, I64 |
| Breast cancer | Self-report | Field ID: 20001 (1002 breast cancer) |
| | Hospital inpatient records | ICD-9: 174; ICD-10: C50 |
| | Death registry | ICD-10: C50 |
| | Cancer registry | ICD-9: 174; ICD-10: C50 |
| Prostate cancer | Self-report | Field ID: 20001 (1044 prostate cancer) |
| | Hospital inpatient records | ICD-9: 1859; ICD-10: C61 |
| | Death registry | ICD-10: C61 |
| | Cancer registry | ICD-9: 1859; ICD-10: C61 |
| Colorectal cancer | Self-report | Field ID: 20001 (1020 large bowel cancer/colorectal cancer, 1021 anal cancer, 1022 colon cancer/sigmoid cancer, 1023 rectal cancer) |
| | Hospital inpatient records | ICD-9: 153, 154; ICD-10: C18, C19, C20 |
| | Death registry | ICD-10: C18, C19, C20 |
| | Cancer registry | ICD-9: 153, 154; ICD-10: C18, C19, C20 |
| Vertebrae fracture | Self-report | Field ID: 20002 (1646 fracture vertebra / crush fracture / vertebral collapse) |
| | Hospital inpatient records | ICD-9: 805, 806; ICD-10: M484, M485, S320, S327 |
| | Death registry | ICD-10: M484, M485, S320, S327 |
| | Cancer registry | ICD-9: 805, 806; ICD-10: M484, M485, S320, S327 |

**Supplementary Table 1. Diagnoses of depression and baseline chronic health conditions (continued)**

| Outcome/health conditions | Source of data | Codes |
|---|---|---|
| Hip fracture | Self-report | Field ID: 20002 (1648 fracture neck of femur / hip) |
| | Hospital inpatient records | ICD-9: 820, 821; ICD-10: S720, S721, S722 |
| | Death registry | ICD-10: S720, S721, S722 |
| | Cancer registry | ICD-9: 820, 821; ICD-10: S720, S721, S722 |
| Dementia | Self-report | Field ID: 20002 (1263 dementia/alzheimers/cognitive impairment) |
| | Hospital inpatient records | ICD-9: 2902, 2903, 2904, 2912, 2941, 3310, 3311, 3312, 3315; ICD-10: A81.0, F00.0, F00.1, F00.2, F00.9, F01.0, F01.1, F01.2, F01.3, F01.8, F01.9, F02.0, F02.0, F02.1, F02.2, F02.3, F02.4, F02.8, F03, F05.1, F10.6, G30.0, G30.1, G30.8, G30.9, G31.0, G31.1, G31.8, I67.3 |
| | Death registry | ICD-10: A81.0, F00.0, F00.1, F00.2, F00.9, F01.0, F01.1, F01.2, F01.3, F01.8, F01.9, F02.0, F02.0, F02.1, F02.2, F02.3, F02.4, F02.8, F03, F05.1, F10.6, G30.0, G30.1, G30.8, G30.9, G31.0, G31.1, G31.8, I67.3 |

### Supplementary Method 2. Definition of covariates

1) **Education level:** divided into higher (college/university degree or other professional qualification), vocational (work-related practical qualifications), upper secondary (second/final stage of secondary education), lower secondary (first stage of secondary education), or other.
2) **Sleep duration:** divided into short (≤ 6 hours), moderate (7-8 hours), and long (>8 hours).
3) **Physical activity:** divided into regular and irregular, with the former defined as at least 150 minutes/week of moderate activity or 75 minutes/week of vigorous activity (or an equivalent combination).
4) **Degree of obesity:** defined according to body mass index (BMI) as thin (< 18.5 $kg/m^2$), normal (18.5-24.9 $kg/m^2$), overweight (25-29.9 $kg/m^2$), or obese (≥ 30.0 $kg/m^2$).
5) **Regular medications taking:** divided into none or yes and the medications involved included cholesterol lowering medication, blood pressure medication, insulin, pain-relief medications (Aspirin, Ibuprofen, Paracetamol, and Codeine), medication for constipation (Laxatives), and medication for heartburn (Ranitidine and Omeprazole).
6) **Healthy plant-based diet index:** made up of 17 food groups, including whole grains, fruits, vegetables, nuts, legumes and vegetarian protein alternatives, tea and coffee, fruit juices, refined grains, potatoes, sugar-sweetened beverages, sweets and desserts, animal fat, dairy, eggs, fish or seafood, meat, and miscellaneous animal-derived foods as previously described [22].

**Supplementary Table 2. The numbers (percentages) of participants with missing covariates**

| Covariates | n | % |
|---|---|---|
| Townsend deprivation index | 125 | 0.1 |
| Education | 357 | 0.3 |
| Race and ethnicity | 373 | 0.3 |
| Smoking history | 247 | 0.2 |
| Sleep duration | 239 | 0.2 |
| Physical activity | 3588 | 3.1 |
| Body mass index | 251 | 0.2 |
| Alcohol intake from non-red drinks | 63 804 | 55.6 |

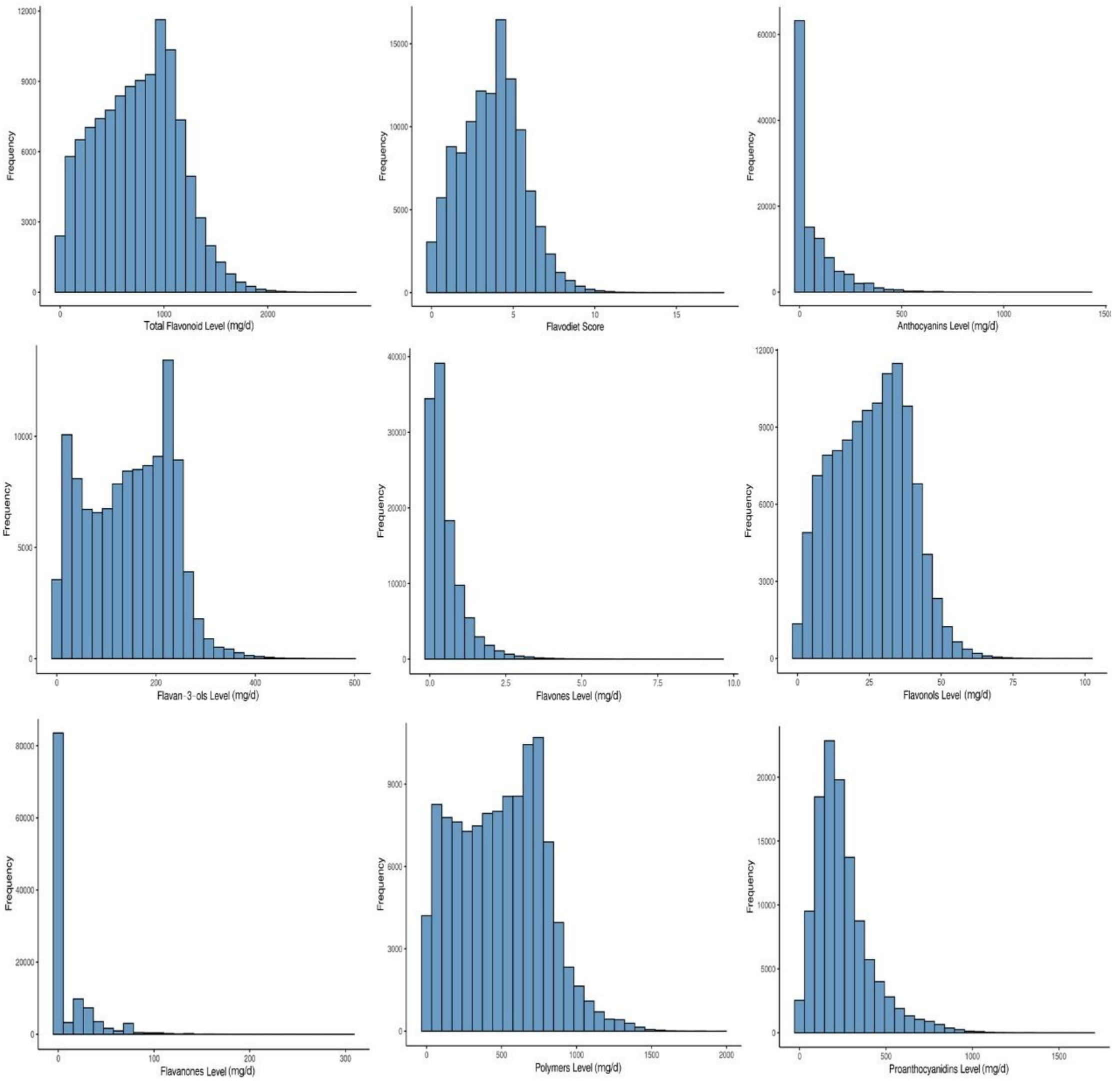


**Supplementary Figure 2. Histogram of intakes of total flavonoid and subclasses**

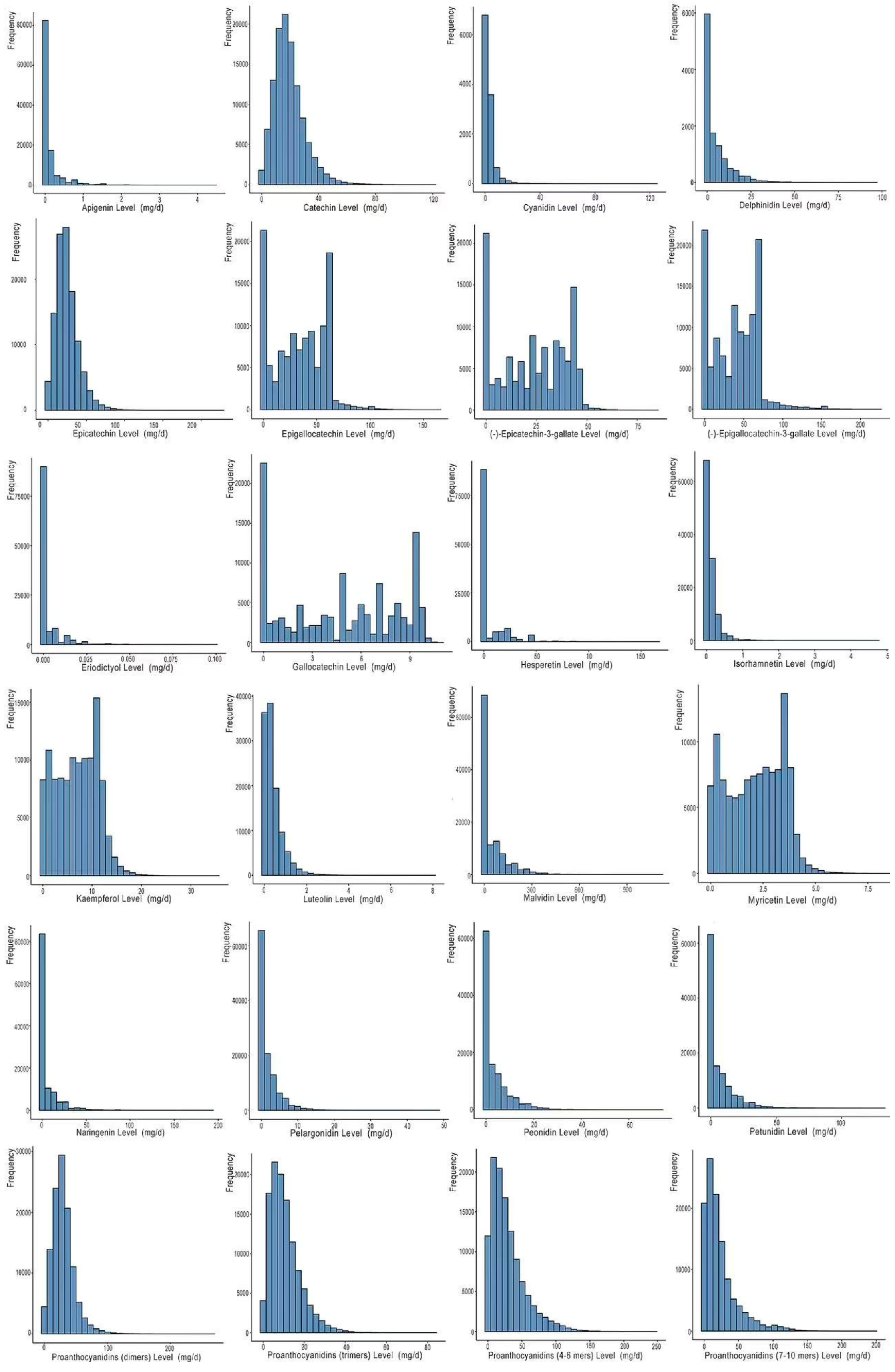


**Supplementary Figure 3. Histogram of intakes of flavonoid compounds**

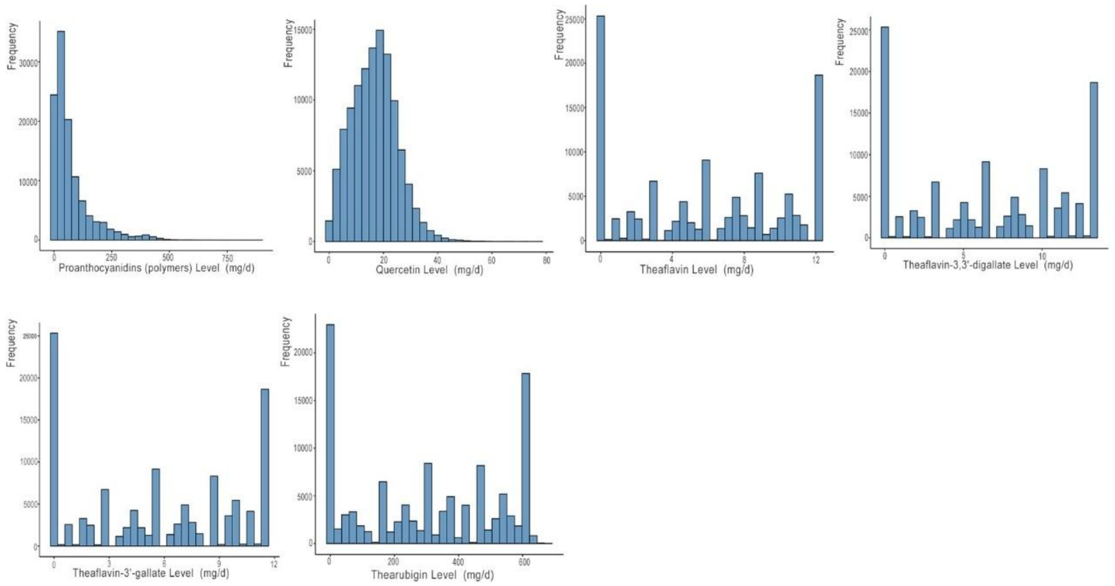


**Supplementary Figure 3. Histogram of intakes of flavonoid compounds (continued)**

**Supplementary Table 3. Baseline characteristics and dietary intakes by incident depression diagnosis**

| Characteristics | Value, median (IQR) | |
|---|---|---|
| | Without depression (n = 111 883) | Incident depression (n = 2965) |
| **Dietary** | | |
| Total flavonoid, mg/d | 760.9 (430.4, 1033.9) | 743.8 (377.0, 1011.3) |
| Anthocyanins, mg/d | 15.5 (1.9, 104.5) | 9.4 (1.5, 63.7) |
| Flavan-3-ols, mg/d | 151.6 (72.6, 217.6) | 146.7 (64.7, 215.3) |
| Flavones, mg/d | 0.3 (0.1, 0.7) | 0.3 (0.1, 0.7) |
| Flavonols, mg/d | 26.3 (15.4, 35.6) | 25.8 (14.4, 34.8) |
| Flavanones, mg/d | 0.9 (0.3, 14.5) | 0.7 (0.2, 11.7) |
| Polymers, mg/d | 497.9 (246.2, 711.5) | 487.9 (228.0, 704.0) |
| Proanthocyanidins, mg/d[a] | 213.1 (140.1, 314.1) | 207.7 (132.5, 310.6) |
| Flavodiet score (tea capped at 4 cups), servings/d[b] | 3.8 (2.3, 5.0) | 3.6 (2.1, 4.8) |
| Tea | 2.2 (0.7, 3.5) | 2.0 (0.5, 3.5) |
| Red wine | 0.0 (0.0, 0.8) | 0.0 (0.0, 0.5) |
| Apples | 0.2 (0.0, 0.7) | 0.2 (0.0, 0.7) |
| Berries | 0.0 (0.0, 0.2) | 0.0 (0.0, 0.2) |
| Grapes | 0.0 (0.0, 0.2) | 0.0 (0.0, 0.2) |
| Grapefruit | 0.0 (0.0, 0.0) | 0.0 (0.0, 0.0) |
| Oranges | 0.0 (0.0, 0.0) | 0.0 (0.0, 0.0) |
| Peppers | 0.0 (0.0, 0.1) | 0.0 (0.0, 0.1) |
| Onions | 0.0 (0.0, 0.2) | 0.0 (0.0, 0.2) |
| Dark chocolate | 0.0 (0.0, 0.0) | 0.0 (0.0, 0.0) |
| Energy, kcal/d | 2009.5 (1719.4, 2331.0) | 1990.1 (1679.9, 2315.7) |
| Fat, g/d | 70.3 (56.2, 86.4) | 69.5 (54.9, 85.9) |
| Alcohol not from red wine, g/d | 5.1 (1.7, 12.0) | 5.1 (1.2, 12.0) |
| hPDI score | 60.8 (57.7, 63.8) | 60.5 (57.1, 63.7) |
| Dietary assessment completion, d | 3.0 (2.0, 4.0) | 3.0 (2.0, 3.0) |
| **Demographic** | | |
| Age, y | 60.4 (53.1, 65.3) | 60.8 (52.9, 65.9) |
| Duration of follow-up, y | 10.5 (10.4, 10.9) | 6.5 (3.9, 8.7) |
| Physical activity, No. (%) | | |
| Irregular | 47 097 (42.1) | 1386 (46.7) |
| Regular | 61 314 (54.8) | 1463 (49.3) |
| Unknown | 3472 (3.1) | 116 (3.9) |
| BMI, kg/m$^2$ | 25.9 (23.5, 28.9) | 26.9 (24.0, 30.6) |
| Sex, No. (%) | | |
| Female | 61 399 (54.9) | 1894 (63.9) |
| Male | 50 484 (45.1) | 1071 (36.1) |
| Race and ethnicity, No. (%) | | |
| Asian | 990 (0.9) | 16 (0.5) |
| Black | 822 (0.7) | 19 (0.6) |

**Supplementary Table 3. Baseline characteristics and dietary intakes by incident depression diagnosis (continued)**

| Characteristics | Value, median (IQR) | |
|---|---|---|
| | Without depression (n = 111 883) | Incident depression (n = 2965) |
| Chinese | 294 (0.3) | 3 (0.1) |
| White | 108 249 (96.8) | 2878 (97.1) |
| Multiracial | 565 (0.5) | 13 (0.4) |
| Other[c] | 604 (0.5) | 22 (0.7) |
| Unknown | 359 (0.3) | 14 (0.5) |
| Townsend deprivation index, No. (%) | | |
| Low | 22 454 (20.1) | 497 (16.8) |
| Moderate | 67 148 (60.0) | 1682 (56.7) |
| High | 22 159 (19.8) | 783 (26.4) |
| Unknown | 122 (0.1) | 3 (0.1) |
| Education level, No. (%) | | |
| Lower secondary | 15 163 (13.6) | 444 (15.0) |
| Upper secondary | 6948 (6.2) | 186 (6.3) |
| Vocational | 11 884 (10.6) | 369 (12.4) |
| Higher | 70 417 (62.9) | 1682 (56.7) |
| Other | 7131 (6.4) | 267 (9.0) |
| Unknown | 340 (0.3) | 17 (0.6) |
| Smoking, No. (%) | | |
| Ever | 47 029 (42.0) | 1498 (50.5) |
| Never | 64 619 (57.8) | 1455 (49.1) |
| Unknown | 235 (0.2) | 12 (0.4) |
| Sleep duration, No. (%) | | |
| >8 | 6385 (5.7) | 239 (8.1) |
| 7-8 | 81 894 (73.2) | 1892 (63.8) |
| ≤ 6 | 23 382 (20.9) | 817 (27.6) |
| Unknown | 222 (0.2) | 17 (0.6) |
| Family history of depression, No. (%) | | |
| Yes | 14 071 (12.6) | 654 (22.1) |
| No | 97 812 (87.4) | 2311 (77.9) |
| No. of medications taken, No. (%) | | |
| 0 | 57 677 (51.6) | 1088 (36.7) |
| 1 | 34 270 (30.6) | 1025 (34.6) |
| 2 | 12 178 (10.9) | 482 (16.3) |
| 3 | 6414 (5.7) | 262 (8.8) |
| >4 | 1344 (1.2) | 108 (3.6) |
| No. of chronic conditions, No. (%) | | |
| 0 | 103 788 (92.8) | 2671 (90.1) |
| 1 | 7839 (7.0) | 280 (9.4) |
| >2 | 256 (0.2) | 14 (0.5) |

Abbreviations: BMI, body mass index; hPDI, healthful plant-based diet index.

[a] Proanthocyanidins are also included in the polymer subclass.

[b] Flavodiet score was calculated by summing intakes (in servings per day) of tea (black and green), red

wine, apples, berries, grapes, oranges, grapefruit, sweet peppers, onions, and dark chocolate.

[c] Other race and ethnicity was self-selected and includes no additional information.

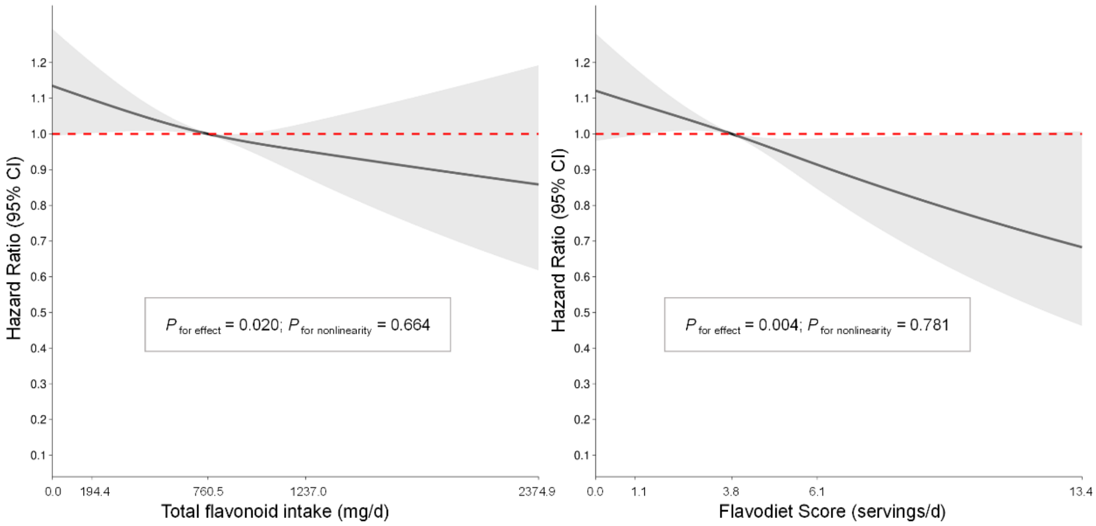


**Supplementary Figure 4. Restricted cubic spline for the association of flavonoid levels with depression risk[a]**

[a] Model adjustment details are provided in the Methods.

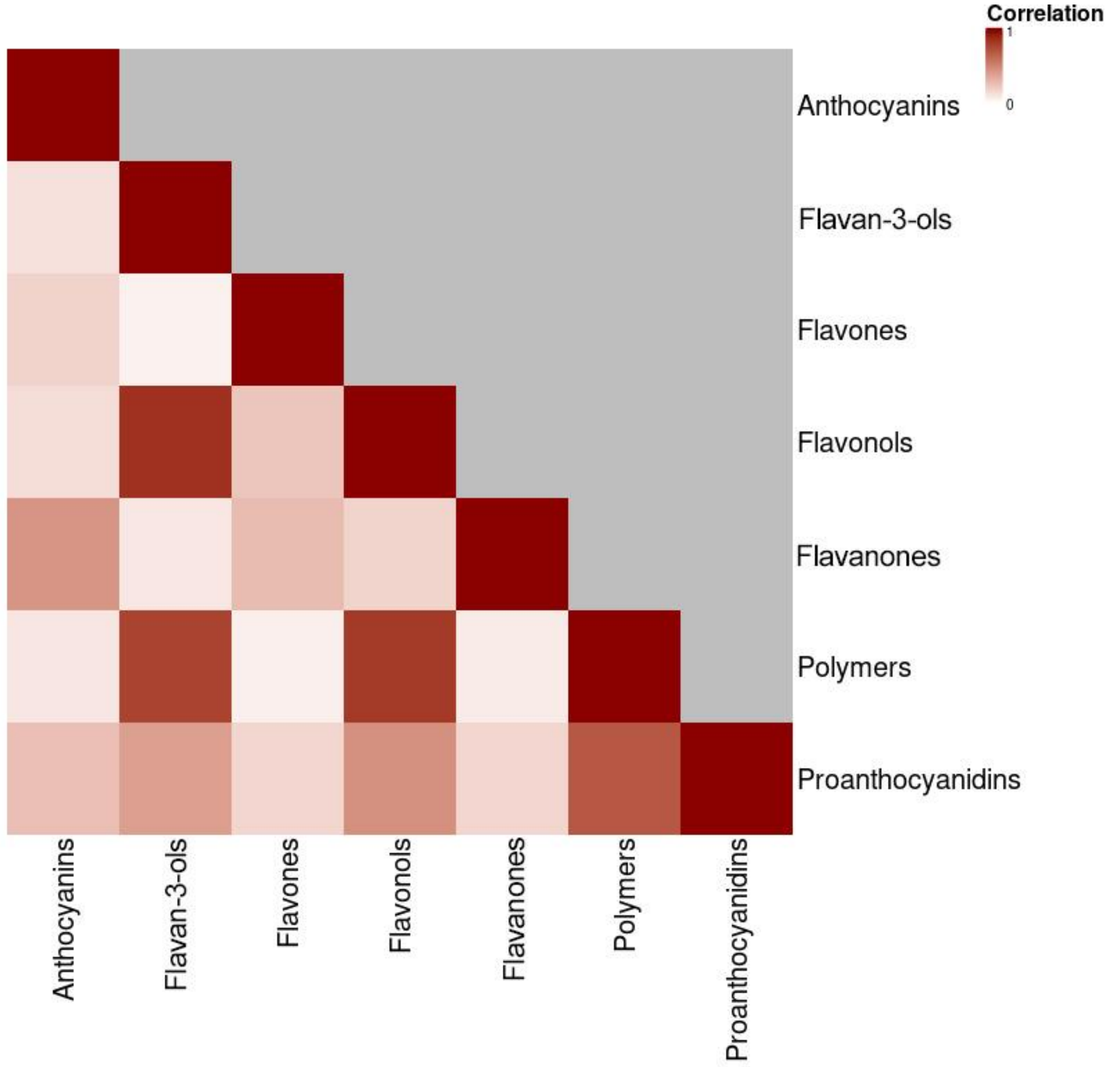


**Supplementary Figure 5. Correlation heatmap for intakes of flavonoid subclasses[a]**

[a] Pearson coefficients after standardization.

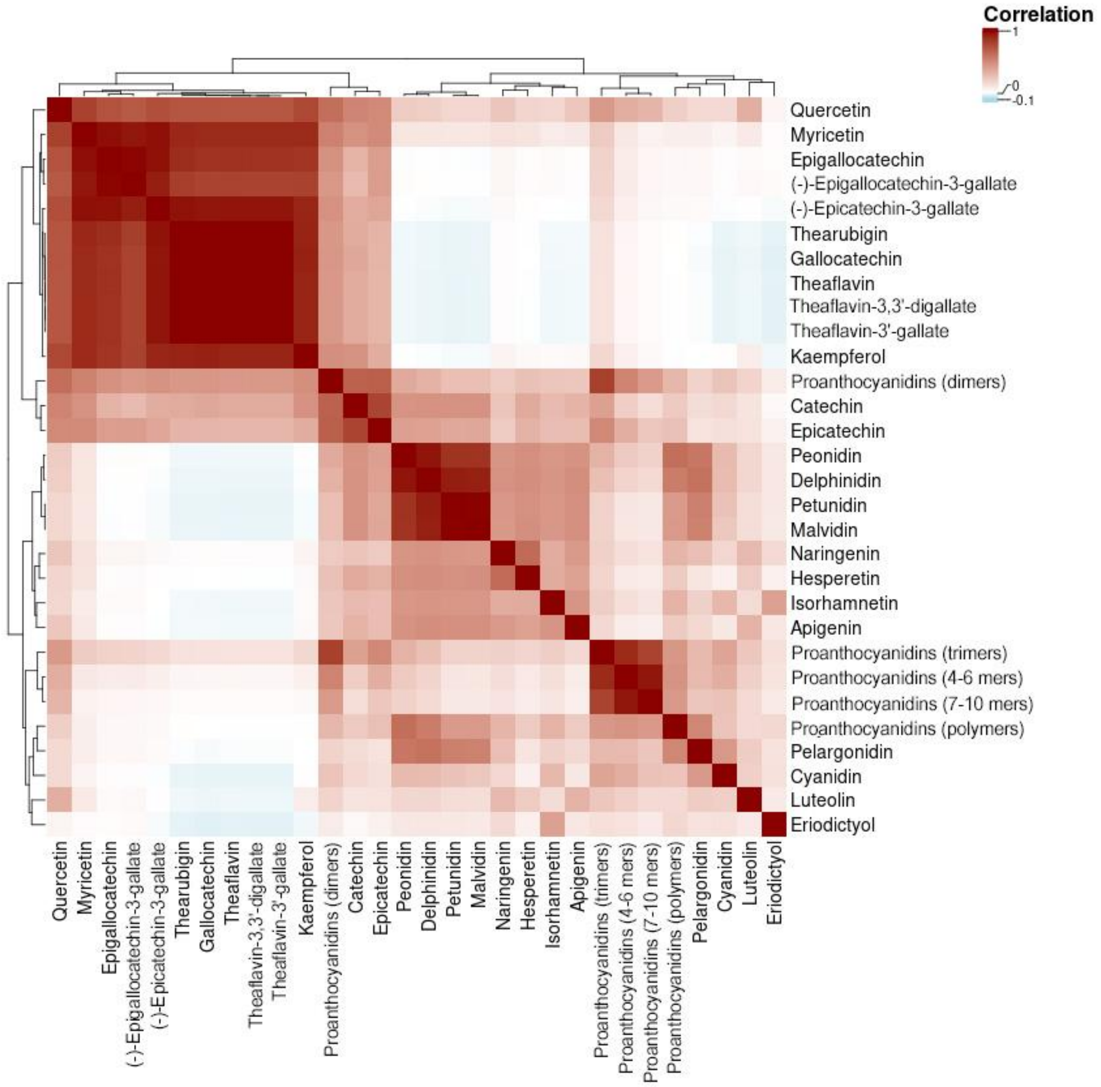


**Supplementary Figure 6. Clustered correlation heatmap for intakes of flavonoid compounds[a]**

[a] Pearson coefficients after standardization.

**Supplementary Table 4. Flavonoid clusters derived from the k-means method**

| Flavonoid compounds | Cluster | Flavonoid subclasses |
|---|---|---|
| Apigenin | 1 | Flavones |
| Hesperetin | 1 | Flavanones |
| Isorhamnetin | 1 | Flavonols |
| Naringenin | 1 | Flavanones |
| Eriodictyol | 2 | Flavanones |
| Luteolin | 2 | Flavones |
| Proanthocyanidins (4-6 mers) | 3 | Polymers, Proanthocyanidins |
| Proanthocyanidins (7-10 mers) | 3 | Polymers, Proanthocyanidins |
| Proanthocyanidins (Trimers) | 3 | Polymers, Proanthocyanidins |
| (-)-Epicatechin-3-gallate | 4 | Flavan-3-ols |
| (-)-Epigallocatechin-3-gallate | 4 | Flavan-3-ols |
| Epigallocatechin | 4 | Flavan-3-ols, Proanthocyanidins |
| Gallocatechin | 4 | Flavan-3-ols, Proanthocyanidins |
| Kaempferol | 4 | Flavonols |
| Myricetin | 4 | Flavonols |
| Quercetin | 4 | Flavonols |
| Theaflavin | 4 | Polymers |
| Theaflavin-3,3'-digallate | 4 | Polymers |
| Theaflavin-3'-gallate | 4 | Polymers |
| Thearubigin | 4 | Polymers |
| Delphinidin | 5 | Anthocyanins |
| Malvidin | 5 | Anthocyanins |
| Peonidin | 5 | Anthocyanins |
| Petunidin | 5 | Anthocyanins |
| Catechin | 6 | Flavan-3-ols, Proanthocyanidins |
| Proanthocyanidins (dimers) | 6 | Polymers, Proanthocyanidins |
| Epicatechin | 6 | Flavan-3-ols, Proanthocyanidins |
| Cyanidin | 7 | Anthocyanins |
| Pelargonidin | 7 | Anthocyanins |
| Proanthocyanidins (polymers) | 7 | Polymers, Proanthocyanidins |

**Supplementary Figure 7. K-means clustering of flavonoid compound intakes**

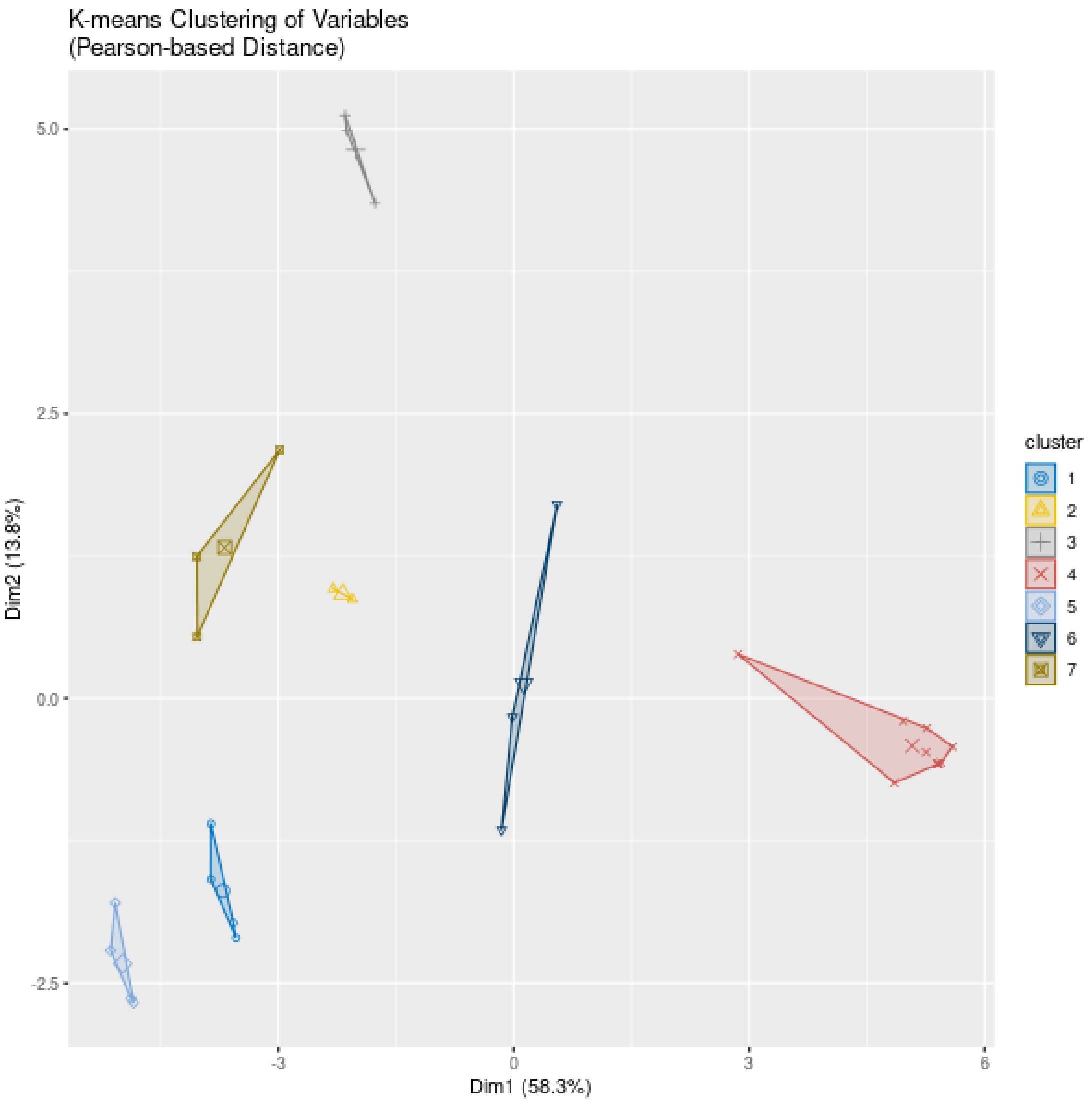

**Supplementary Table 5. The top two food items with the highest content of each compound within Cluster1, Cluster 5, and Cluster 6 from the k-means[a,b]**

| | Cluster 1 (%) | | | | Cluster 5 (%) | | | | Cluster 6 (%) | | |
|---|---|---|---|---|---|---|---|---|---|---|---|
| | Apigenin | Hesperetin | Isorhamnetin | Naringenin | Delphinidin | Malvidin | Peonidin | Petunidin | Catechin | Proanthocyanidins (Dimers) | Epicatechin |
| Black tea | 0 | 0 | 0 | 0 | 0 | 0 | 0 | 0 | **33.3** | **40.6** | **37.7** |
| Green tea | 0 | 0 | 0 | 0 | 0 | 0 | 0 | 0 | 0 | 1.9 | 3.3 |
| Red wine | **35.8** | **54** | **31** | **23.2** | **39.4** | **52.6** | **39.6** | **50.8** | 15.5 | 11.9 | **15.4** |
| Apple | 0 | 0 | 0 | 0 | 20.4 | 0 | 20.2 | 0 | 3.9 | **16.5** | 13.1 |
| Berries | 0 | 0 | 0 | 7.4 | **26.2** | **28.3** | **22.4** | **32.3** | 4.7 | 2.8 | 1.6 |
| Grapes | 0 | 0 | 0 | 0 | 12.9 | 19.1 | 11.7 | 16.9 | 4.9 | 1.6 | 3.6 |
| Grapefruit | 0 | 6.7 | 0 | 5.4 | 0 | 0 | 0 | 0 | 0 | 0 | 0 |
| Orange | 0 | **39.3** | 0 | 20.5 | 0 | 0 | 0 | 0 | 0 | 0 | 0 |
| Dark chocolate | 0 | 0 | 0 | 0 | 0 | 0 | 0 | 0 | 1.5 | 2.5 | 2.8 |
| Bran cereal | 0 | 0 | 0 | 0 | 0 | 0 | 0 | 0 | 0 | 3.3 | 0 |
| Olive oil | 2 | 0 | 0 | 0 | 0 | 0 | 0 | 0 | 0 | 0 | 0 |
| Avocado | 0 | 0 | 0 | 0 | 0 | 0 | 0 | 0 | 0 | 0.2 | 0.1 |
| Broad bean | 0 | 0 | 0 | 0 | 0 | 0 | 0 | 0 | 1 | 2.6 | 0.9 |
| Cabbage kale | 8.5 | 0 | 19.6 | 0 | 0 | 0 | 0 | 0 | 0 | 0 | 0 |
| Celery | 18.1 | 0 | 0 | 0 | 0 | 0 | 0 | 0 | 0 | 0 | 0 |
| Garlic | 5 | 0 | 0 | 0 | 0 | 0 | 0 | 0 | 0 | 0 | 0 |
| Lettuce | **28.3** | 0 | 0 | 0 | 0 | 0 | 0 | 0 | 0 | 0 | 0 |
| Sprouts | 0 | 0 | 0 | 3.5 | 0 | 0 | 0 | 0 | 0 | 0 | 0 |
| Fresh tomato | 0 | 0 | 0 | **36.9** | 0 | 0 | 0 | 0 | 0 | 0 | 0 |

**Supplementary Table 5. The top two food items with the highest content of each compound within Cluster1, Cluster 5, and Cluster 6 from the k-means[a,b] (continued)**

| | Cluster 1 (%) | | | | Cluster 5 (%) | | | | Cluster 6 (%) | | |
|---|---|---|---|---|---|---|---|---|---|---|---|
| | Apigenin | Hesperetin | Isorhamnetin | Naringenin | Delphinidin | Malvidin | Peonidin | Petunidin | Catechin | Proanthocyanidins (Dimers) | Epicatechin |
| Banana | 0 | 0 | 0 | 0 | 0 | 0 | 0 | 0 | **16.6** | 1.8 | 0.8 |
| Cherry | 0 | 0 | 7.5 | 0 | 0 | 0 | 1.7 | 0 | 0.2 | 0 | 0.4 |
| Mango | 2.3 | 0 | 0 | 0 | 1.2 | 0 | 0 | 0 | 0.4 | 0.3 | 0 |
| Peach nectarine | 0 | 0 | 0 | 0 | 0 | 0 | 0 | 0 | 1.6 | 1.8 | 1 |
| Pear | 0 | 0 | **22.6** | 0 | 0 | 0 | 0 | 0 | 0.3 | 1 | 1.9 |
| Plum | 0 | 0 | 0 | 0 | 0 | 0 | 4.4 | 0 | 0.8 | 3.4 | 0.8 |
| Salted peanuts | 0 | 0 | 0 | 0 | 0 | 0 | 0 | 0 | 0 | 0.3 | 0 |
| Unsalted peanuts | 0 | 0 | 0 | 0 | 0 | 0 | 0 | 0 | 0 | 0.3 | 0 |
| Salted nuts | 0 | 0 | 5.1 | 0.9 | 0 | 0 | 0 | 0 | 0.1 | 0.3 | 0.1 |
| Unsalted nuts | 0 | 0 | 14.2 | 2.2 | 0 | 0 | 0 | 0 | 0.4 | 0.8 | 0.2 |
| Seeds | 0 | 0 | 0 | 0 | 0 | 0 | 0 | 0 | 0.1 | 0 | 0 |
| Milk chocolate | 0 | 0 | 0 | 0 | 0 | 0 | 0 | 0 | 0.6 | 2.2 | 0.8 |
| Chocolate confectionery | 0 | 0 | 0 | 0 | 0 | 0 | 0 | 0 | 6.7 | 0 | 12.5 |
| Filtered coffee | 0 | 0 | 0 | 0 | 0 | 0 | 0 | 0 | 0 | 0 | 0.7 |
| Beer cider | 0 | 0 | 0 | 0 | 0 | 0 | 0 | 0 | 7.5 | 4 | 2.4 |
| Total | 100 | 100 | 100 | 100 | 100 | 100 | 100 | 100 | 100 | 100 | 100 |

[a] Food items with zero content for all compounds listed in the table were not shown.

[b] Values representing the top two highest concentrations for each compound are demonstrated in bold.

**Supplementary Table 6. Risk of depression by quintiles of flavonoid intake levels in participants aged more than 60 years[a]**

| Intake component | Quintile 1 (lowest) | Quintile 2 | Quintile 3 | Quintile 4 | Quintile 5 (highest) | P for trend | HR per SD (95% CI) |
|---|---|---|---|---|---|---|---|
| Total flavonoid, mg/d | | | | | | | |
| Intake, median (IQR) | 232.0 (130.6, 318.6) | 551.6 (477.8, 617.5) | 805.5 (745.1, 862.5) | 1012.0 (966.6, 1058.4) | 1261.7 (1175.8, 1396.8) | | |
| Case/N | 349/11 849 | 287/11 849 | 316/11 849 | 323/11 849 | 282/11 849 | 0.108 | 0.96 (0.91, 1.01) |
| HR (95% CI) | 1 (reference) | 0.85 (0.73, 0.99) | 0.94 (0.81, 1.1) | 0.95 (0.81, 1.10) | 0.82 (0.70, 0.97) | | |
| Flavodiet Score, points[b] | | | | | | | |
| Intake, median (IQR) | 1.3 (0.8, 1.8) | 2.9 (2.5, 3.1) | 4 (3.7, 4.2) | 4.9 (4.6, 5.1) | 6.2 (5.8, 7.0) | | |
| Case/N | 342/11 856 | 289/11 881 | 339/11 884 | 307/11 797 | 280/11 827 | 0.098 | 0.94 (0.90, 1.00) |
| HR (95% CI) | 1 (reference) | 0.87 (0.74, 1.01) | 1.01 (0.87, 1.18) | 0.93 (0.79, 1.09) | 0.83 (0.70, 0.98) | | |
| Anthocyanins, mg/d | | | | | | | |
| Intake, median (IQR) | 0.6 (0.0, 1.2) | 4.2 (2.7, 6.7) | 21.5 (13.9, 33.6) | 87.1 (69.4, 113.4) | 223.7 (171.2, 304.4) | | |
| Case/N | 351/12 124 | 368/11 575 | 344/11 848 | 251/11 849 | 243/11 849 | **<0.001** | 0.92 (0.88, 0.97) |
| HR (95% CI) | 1 (reference) | 1.11 (0.96, 1.29) | 0.99 (0.85, 1.15) | 0.77 (0.65, 0.91) | 0.72 (0.61, 0.85) | | |
| Flavan-3-ols, mg/d | | | | | | | |
| Intake, median (IQR) | 29.3 (16.2, 44.4) | 99.4 (81.4, 116.4) | 159 (145.2, 172) | 209.8 (197.4, 220.1) | 246.7 (235.8, 266.5) | | |
| Case/N | 336/11 849 | 292/11 849 | 316/11 849 | 323/11 849 | 290/11 849 | 0.515 | 0.98 (0.93, 1.03) |
| HR (95% CI) | 1 (reference) | 0.91 (0.78, 1.07) | 0.99 (0.85, 1.15) | 0.99 (0.85, 1.16) | 0.90 (0.76, 1.05) | | |
| Flavones, mg/d | | | | | | | |
| Intake, median (IQR) | 0 (0, 0.1) | 0.2 (0.1, 0.2) | 0.3 (0.3, 0.4) | 0.6 (0.5, 0.7) | 1.3 (1.0, 1.7) | | |
| Case/N | 343/11 850 | 319/11 848 | 293/11 849 | 270/11 849 | 332/11 849 | 0.673 | 1.01 (0.96, 1.06) |
| HR (95% CI) | 1 (reference) | 0.93 (0.8, 1.09) | 0.87 (0.74, 1.02) | 0.8 (0.68, 0.94) | 0.94 (0.80, 1.11) | | |
| Flavonols, mg/d | | | | | | | |

**Supplementary Table 6. Risk of depression by quintiles of flavonoid intake levels in participants aged more than 60 years[a] (continued)**

| Intake component | Quintile 1 (lowest) | Quintile 2 | Quintile 3 | Quintile 4 | Quintile 5 (highest) | P for trend | HR per SD (95% CI) |
|---|---|---|---|---|---|---|---|
| Intake, median (IQR) | 8.6 (5.6, 11.4) | 19 (16.6, 21.2) | 27.4 (25.4, 29.4) | 34.6 (32.9, 36.2) | 42.5 (40.0, 46.6) | | |
| Case/N | 330/11 849 | 299/11 849 | 301/11 849 | 329/11 849 | 298/11 849 | 0.717 | 0.99 (0.94, 1.04) |
| HR (95% CI) | 1 (reference) | 0.93 (0.8, 1.09) | 0.94 (0.80, 1.10) | 1.03 (0.88, 1.2) | 0.92 (0.78, 1.09) | | |
| Flavanones, mg/g | | | | | | | |
| Intake, median (IQR) | 0.0 (0.0, 0.1) | 0.5 (0.3, 0.6) | 1.1 (0.9, 1.3) | 11.4 (2.3, 18.1) | 44.2 (34.4, 68.1) | | |
| Case/N | 373/11 849 | 319/11 849 | 274/11 849 | 296/11 849 | 295/11 849 | **0.005** | 0.94 (0.89, 0.98) |
| HR (95% CI) | 1 (reference) | 0.86 (0.74, 1.00) | 0.74 (0.63, 0.87) | 0.82 (0.70, 0.96) | 0.79 (0.68, 0.93) | | |
| Polymers, mg/d | | | | | | | |
| Intake, median (IQR) | 111.8 (58.7, 165.2) | 332.4 (275, 383.7) | 524.6 (479.1, 570.8) | 694.9 (659.1, 727.8) | 861.1 (801.0, 970.1) | | |
| Case/N | 331/11 849 | 304/11 849 | 302/11 849 | 319/11 849 | 301/11 849 | 0.496 | 0.99 (0.94, 1.05) |
| HR (95% CI) | 1 (reference) | 0.94 (0.80, 1.10) | 0.95 (0.81, 1.11) | 0.98 (0.84, 1.14) | 0.92 (0.78, 1.08) | | |
| Proanthocyanidins, mg/d[c] | | | | | | | |
| Intake, median (IQR) | 95.8 (63.5, 117.9) | 165 (150.4, 179.5) | 223.5 (208.7, 239.4) | 299.0 (276.5, 327.0) | 475.3 (408.6, 591.7) | | |
| Case/N | 323/11 849 | 321/11 849 | 320/11 849 | 285/11 849 | 308/11 849 | 0.466 | 0.98 (0.93, 1.03) |
| HR (95% CI) | 1 (reference) | 1.03 (0.88, 1.21) | 1.02 (0.87, 1.19) | 0.91 (0.77, 1.07) | 0.98 (0.83, 1.15) | | |

[a] Total number of participants, 59 245; number of participants with incident depression, 1557.

[b] Flavodiet score was calculated by summing intakes (in servings per day) of tea (black and green), red wine, apples, berries, grapes, oranges, grapefruit, sweet peppers, onions, and dark chocolate.

[c] Proanthocyanidins are also included in the polymer subclass. Model adjustment details are provided in the Methods.

**Supplementary Table 7. Risk of depression by quintiles of flavonoid intake levels in participants excluding cases diagnosed within 2 years of follow-up[a]**

| Intake component | Quintile 1 (lowest) | Quintile 2 | Quintile 3 | Quintile 4 | Quintile 5 (highest) | P for trend | HR per SD (95% CI) |
|---|---|---|---|---|---|---|---|
| Total flavonoid, mg/d | | | | | | | |
| Intake, median (IQR) | 194.5 (108.2, 276.1) | 501.6 (429.2, 572.2) | 760.5 (700.4, 822.3) | 985.5 (937.1, 1033.3) | 1237.0 (1150.5, 1375.8) | **0.015** | 0.94 (0.90, 0.98) |
| Case/N | 603/22 904 | 501/22 903 | 526/22 903 | 524/22 903 | 479/22 903 | | |
| HR (95% CI) | 1 (reference) | 0.87 (0.77, 0.98) | 0.92 (0.82, 1.04) | 0.90 (0.80, 1.01) | 0.83 (0.73, 0.94) | | |
| Flavodiet Score, points[b] | | | | | | | |
| Intake, median (IQR) | 1.1 (0.6, 1.5) | 2.6 (2.3, 3.0) | 3.8 (3.5, 4.0) | 4.8 (4.5, 5.0) | 6.1 (5.6, 6.9) | **<0.001** | 0.93 (0.89, 0.97) |
| Case/N | 610/22 913 | 537/24 056 | 542/22 800 | 499/21 925 | 445/22 822 | | |
| HR (95% CI) | 1 (reference) | 0.86 (0.77, 0.97) | 0.92 (0.82, 1.03) | 0.89 (0.79, 1.00) | 0.75 (0.66, 0.85) | | |
| Anthocyanins, mg/d | | | | | | | |
| Intake, median (IQR) | 0.3 (0.0, 0.9) | 2.8 (1.9, 4.5) | 15.4 (11.2, 24.2) | 82.0 (58.7, 102.5) | 211.8 (165.6, 292.5) | **<0.001** | 0.90 (0.87, 0.93) |
| Case/N | 633/23 381 | 599/22 427 | 582/22 902 | 450/22 903 | 369/22 903 | | |
| HR (95% CI) | 1 (reference) | 1.03 (0.91, 1.15) | 0.94 (0.83, 1.06) | 0.79 (0.69, 0.89) | 0.61 (0.54, 0.70) | | |
| Flavan-3-ols, mg/d | | | | | | | |
| Intake, median (IQR) | 26.2 (14.6, 38.9) | 90.2 (72.4, 107.9) | 151.5 (138.0, 165.6) | 205.4 (192.4, 217.6) | 245.5 (234.4, 265.7) | 0.068 | 0.96 (0.92, 1.00) |
| Case/N | 590/22 904 | 514/22 903 | 507/22 903 | 538/22 914 | 484/22 892 | | |
| HR (95% CI) | 1 (reference) | 0.92 (0.82, 1.04) | 0.91 (0.81, 1.03) | 0.95 (0.84, 1.07) | 0.86 (0.76, 0.98) | | |
| Flavones, mg/d | | | | | | | |
| Intake, median (IQR) | 0.0 (0.0, 0.1) | 0.2 (0.1, 0.2) | 0.3 (0.3, 0.4) | 0.6 (0.5, 0.7) | 1.2 (1.0, 1.7) | 0.481 | 1.00 (0.96, 1.04) |
| Case/N | 615/22 924 | 490/22 883 | 500/22 903 | 485/22 903 | 543/22 903 | | |
| HR (95% CI) | 1 (reference) | 0.83 (0.74, 0.94) | 0.86 (0.76, 0.97) | 0.83 (0.74, 0.94) | 0.89 (0.78, 1.01) | | |
| Flavonols, mg/d | | | | | | | |
| Intake, median (IQR) | 7.8 (5.0, 10.4) | 17.7 (15.4, 20.0) | 26.3 (24.3, 28.3) | 33.8 (32.1, 35.5) | 42.0 (39.4, 46.0) | 0.199 | 0.96 (0.93, 1.00) |
| Case/N | 590/22 904 | 499/22 903 | 505/22 903 | 538/22 903 | 501/22 903 | | |
| HR (95% CI) | 1 (reference) | 0.88 (0.78, 0.99) | 0.90 (0.80, 1.02) | 0.95 (0.84, 1.07) | 0.88 (0.78, 1.00) | | |

**Supplementary Table 7. Risk of depression by quintiles of flavonoid intake levels in participants excluding cases diagnosed within 2 years of follow-up (continued)**

| Intake component | Quintile 1 (lowest) | Quintile 2 | Quintile 3 | Quintile 4 | Quintile 5 (highest) | P for trend | HR per SD (95% CI) |
|---|---|---|---|---|---|---|---|
| Flavanones, mg/d | | | | | | | |
| Intake, median (IQR) | 0.0 (0.0, 0.1) | 0.4 (0.3, 0.5) | 0.9 (0.7, 1.1) | 2.6 (1.8, 14.7) | 35.9 (32.7, 58.9) | **<0.001** | 0.93 (0.89, 0.96) |
| Case/N | 640/23 017 | 576/23 122 | 449/22 594 | 456/23 079 | 512/22 704 | | |
| HR (95% CI) | 1 (reference) | 0.91 (0.81, 1.02) | 0.73 (0.65, 0.83) | 0.73 (0.64, 0.83) | 0.81 (0.72, 0.92) | | |
| Polymers, mg/d | | | | | | | |
| Intake, median (IQR) | 93.7 (46.8, 141.2) | 298.7 (245.7, 351.7) | 497.7 (449.6, 543.1) | 682.8 (635.2, 711.4) | 842.2 (785.4, 952.1) | 0.378 | 0.98 (0.94, 1.02) |
| Case/N | 589/22 905 | 497/22 902 | 494/22 903 | 536/22 907 | 517/22 899 | | |
| HR (95% CI) | 1 (reference) | 0.88 (0.78, 0.99) | 0.89 (0.79, 1.00) | 0.94 (0.84, 1.06) | 0.91 (0.81, 1.03) | | |
| Proanthocyanidins, mg/d[c] | | | | | | | |
| Intake, median (IQR) | 84.3 (54.2, 107.3) | 154.7 (140.0, 169.3) | 213.0 (198.1, 228.7) | 287.3 (265.6, 314) | 459.8 (393.4, 576.5) | 0.254 | 0.98 (0.94, 1.02) |
| Case/N | 586/22 904 | 545/22 903 | 487/22 903 | 509/22 903 | 506/22 903 | | |
| HR (95% CI) | 1 (reference) | 0.97 (0.86, 1.09) | 0.88 (0.78, 1.00) | 0.93 (0.82, 1.05) | 0.92 (0.81, 1.05) | | |

[a] Total number of participants, 114 516; number of participants with incident depression, 2633.

[b] Flavodiet score was calculated by summing intakes (in servings per day) of tea (black and green), red wine, apples, berries, grapes, oranges, grapefruit, sweet peppers, onions, and dark chocolate.

[c] Proanthocyanidins are also included in the polymer subclass. Model adjustment details are provided in the Methods.

**Supplementary Table 8. Risk of depression by quintiles of flavonoid intake levels in participants with at least 5 years follow-up[a]**

| Intake component | Quintile 1 (lowest) | Quintile 2 | Quintile 3 | Quintile 4 | Quintile 5 (highest) | P for trend | HR per SD (95% CI) |
|---|---|---|---|---|---|---|---|
| Total flavonoid, mg/d | | | | | | | |
| Intake, median (IQR) | 195.6 (109.2, 277.2) | 502.3 (430.2, 572.6) | 760.8 (700.8, 822.7) | 985.5 (937.2, 1033.3) | 1237.3 (1150.5, 1376.4) | | |
| Case/N | 434/22 361 | 377/22 360 | 386/22 361 | 399/22 360 | 338/22 361 | **0.006** | 0.93 (0.89, 0.98) |
| HR (95% CI) | 1 (reference) | 0.89 (0.78, 1.03) | 0.91 (0.80, 1.05) | 0.92 (0.80, 1.06) | 0.78 (0.67, 0.90) | | |
| Flavodiet Score, points[b] | | | | | | | |
| Intake, median (IQR) | 1.2 (0.6, 1.7) | 2.7 (2.4, 3) | 3.8 (3.5, 4.0) | 4.8 (4.5, 5.0) | 6.1 (5.6, 6.9) | | |
| Case/N | 463/24 362 | 370/21 451 | 412/22 259 | 367/21 427 | 322/22 304 | **0.001** | 0.92 (0.88, 0.97) |
| HR (95% CI) | 1 (reference) | 0.92 (0.80, 1.06) | 0.98 (0.85, 1.12) | 0.91 (0.79, 1.05) | 0.74 (0.64, 0.87) | | |
| Anthocyanins, mg/d | | | | | | | |
| Intake, median (IQR) | 0.3 (0.0, 0.9) | 2.9 (1.9, 4.5) | 15.5 (11.2, 24.5) | 82.0 (59.2, 103.6) | 212.1 (165.7, 292.4) | | |
| Case/N | 462/22 733 | 423/21 988 | 435/22 445 | 340/22 276 | 274/22 361 | **<0.001** | 0.89 (0.85, 0.92) |
| HR (95% CI) | 1 (reference) | 0.96 (0.84, 1.10) | 0.93 (0.81, 1.06) | 0.78 (0.68, 0.91) | 0.59 (0.51, 0.69) | | |
| Flavan-3-ols, mg/d | | | | | | | |
| Intake, median (IQR) | 26.3 (14.7, 39.0) | 90.3 (72.6, 108.0) | 151.5 (138.0, 165.6) | 205.4 (192.3, 217.5) | 245.5 (234.4, 265.7) | | |
| Case/N | 425/22 361 | 385/22 363 | 374/22 358 | 395/22 360 | 355/22 361 | 0.069 | 0.96 (0.91, 1.00) |
| HR (95% CI) | 1 (reference) | 0.94 (0.82, 1.09) | 0.91 (0.79, 1.05) | 0.95 (0.82, 1.09) | 0.86 (0.74, 0.99) | | |
| Flavones, mg/d | | | | | | | |
| Intake, median (IQR) | 0.0 (0.0, 0.1) | 0.2 (0.1, 0.2) | 0.3 (0.3, 0.4) | 0.6 (0.5, 0.7) | 1.2 (1.0, 1.7) | | |
| Case/N | 459/22 362 | 349/22 360 | 368/22 360 | 351/22 360 | 407/22 361 | 0.685 | 1.00 (0.96, 1.05) |
| HR (95% CI) | 1 (reference) | 0.78 (0.67, 0.89) | 0.82 (0.71, 0.94) | 0.78 (0.68, 0.91) | 0.88 (0.76, 1.01) | | |
| Flavonols, mg/d | | | | | | | |

| Intake component | Quintile 1 (lowest) | Quintile 2 | Quintile 3 | Quintile 4 | Quintile 5 (highest) | P for trend | HR per SD (95% CI) |
|---|---|---|---|---|---|---|---|
| Intake, median (IQR) | 7.8 (5.0, 10.5) | 17.8 (15.4, 20.0) | 26.3 (24.3, 28.3) | 33.8 (32.1, 35.5) | 42.0 (39.4, 46.0) | 0.125 | 0.95 (0.91, 1.00) |
| Case/N | 430/22 361 | 367/22 360 | 373/22 361 | 406/22 360 | 358/22 361 | | |
| HR (95% CI) | 1 (reference) | 0.87 (0.76, 1.00) | 0.89 (0.77, 1.02) | 0.96 (0.84, 1.10) | 0.84 (0.72, 0.97) | | |
| Flavanones, mg/g | | | | | | | |
| Intake, median (IQR) | 0.0 (0.0, 0.1) | 0.4 (0.3, 0.5) | 0.9 (0.7, 1.1) | 2.6 (1.8, 14.7) | 35.9 (32.2, 59.2) | **<0.001** | 0.93 (0.89, 0.97) |
| Case/N | 461/22 368 | 412/22 581 | 334/22 133 | 343/22 557 | 384/22 164 | | |
| HR (95% CI) | 1 (reference) | 0.89 (0.77, 1.02) | 0.73 (0.63, 0.84) | 0.73 (0.63, 0.84) | 0.80 (0.70, 0.92) | | |
| Polymers, mg/d | | | | | | | |
| Intake, median (IQR) | 94.0 (47.1, 141.8) | 299.2 (246.3, 352.3) | 497.6 (449.6, 543.1) | 682.8 (635.2, 711.5) | 842.1 (785.4, 952.4) | 0.261 | 0.97 (0.93, 1.02) |
| Case/N | 427/22 361 | 368/22 360 | 365/22 361 | 396/22 365 | 378/22 356 | | |
| HR (95% CI) | 1 (reference) | 0.88 (0.77, 1.01) | 0.88 (0.77, 1.02) | 0.94 (0.81, 1.07) | 0.89 (0.77, 1.03) | | |
| Proanthocyanidins, mg/d[c] | | | | | | | |
| Intake, median (IQR) | 84.5 (54.3, 107.5) | 154.8 (140.0, 169.4) | 213.1 (198.2, 228.8) | 287.4 (265.7, 314.1) | 459.9 (393.5, 577.0) | **0.041** | 0.96 (0.91, 1.01) |
| Case/N | 434/22 361 | 410/22 360 | 356/22 361 | 366/22 360 | 368/22 361 | | |
| HR (95% CI) | 1 (reference) | 0.96 (0.84, 1.1) | 0.83 (0.72, 0.96) | 0.86 (0.74, 0.99) | 0.86 (0.74, 1.00) | | |

[a] Total number of participants, 111 803; number of participants with incident depression, 1934.

[b] Flavodiet score was calculated by summing intakes (in servings per day) of tea (black and green), red wine, apples, berries, grapes, oranges, grapefruit, sweet peppers, onions, and dark chocolate.

[c] Proanthocyanidins are also included in the polymer subclass. Model adjustment details are provided in the Methods.

**Supplementary Table 9. Risk of depression by quintiles of flavonoid intake levels in participants living in areas of high deprivation or with low levels of education[a]**

| Intake component | Quintile 1 (lowest) | Quintile 2 | Quintile 3 | Quintile 4 | Quintile 5 (highest) | P for trend | HR per SD (95% CI) |
|---|---|---|---|---|---|---|---|
| Total flavonoid, mg/d | | | | | | | |
| Intake, median (IQR) | 183.5 (101.1, 264.2) | 487.0 (414.6, 556.9) | 748.4 (687.9, 811.9) | 976.3 (929.8, 1024.0) | 1226.5 (1140.8, 1366.5) | **0.010** | 0.94 (0.90, 0.98) |
| Case/N | 502/15 729 | 439/15 728 | 424/15 728 | 448/15 728 | 387/15 728 | | |
| HR (95% CI) | 1 (reference) | 0.92 (0.81, 1.05) | 0.9 (0.79, 1.03) | 0.93 (0.82, 1.06) | 0.81 (0.71, 0.93) | | |
| Flavodiet Score, points[b] | | | | | | | |
| Intake, median (IQR) | 1.0 (0.5, 1.5) | 2.5 (2.2, 2.9) | 3.7 (3.5, 4.0) | 4.7 (4.5, 5.0) | 6.1 (5.6, 6.9) | **<0.001** | 0.91 (0.87, 0.95) |
| Case/N | 518/15 868 | 435/15 589 | 461/15 799 | 445/16 260 | 341/15 125 | | |
| HR (95% CI) | 1 (reference) | 0.89 (0.79, 1.02) | 0.93 (0.82, 1.06) | 0.88 (0.77, 1.00) | 0.71 (0.62, 0.83) | | |
| Anthocyanins, mg/d | | | | | | | |
| Intake, median (IQR) | 0.1 (0.0, 0.8) | 2.5 (1.7, 3.7) | 13.2 (9.1, 21.0) | 78.1 (55.0, 96.2) | 211.0 (164.9, 295.2) | **<0.001** | 0.90 (0.86, 0.93) |
| Case/N | 526/15 769 | 510/15 716 | 482/15 700 | 383/15 774 | 299/15 682 | | |
| HR (95% CI) | 1 (reference) | 1.01 (0.89, 1.15) | 0.94 (0.82, 1.07) | 0.80 (0.69, 0.91) | 0.60 (0.52, 0.69) | | |
| Flavan-3-ols, mg/d | | | | | | | |
| Intake, median (IQR) | 25.2 (13.9, 37.4) | 87.6 (69.5, 105.5) | 149.7 (136.0, 164.1) | 204.3 (191.2, 217) | 245.2 (233.9, 265.9) | **0.021** | 0.95 (0.91, 0.99) |
| Case/N | 500/15 729 | 438/15 728 | 418/15 728 | 457/15 728 | 387/15 728 | | |
| HR (95% CI) | 1 (reference) | 0.93 (0.82, 1.06) | 0.90 (0.79, 1.02) | 0.96 (0.84, 1.09) | 0.82 (0.71, 0.94) | | |
| Flavones, mg/d | | | | | | | |
| Intake, median (IQR) | 0.0 (0.0, 0.1) | 0.2 (0.1, 0.2) | 0.3 (0.3, 0.4) | 0.6 (0.5, 0.7) | 1.2 (1.0, 1.7) | 0.267 | 0.99 (0.95, 1.04) |
| Case/N | 528/15 729 | 418/15 729 | 402/15 727 | 414/15 728 | 438/15 728 | | |
| HR (95% CI) | 1 (reference) | 0.83 (0.73, 0.95) | 0.81 (0.71, 0.93) | 0.85 (0.74, 0.97) | 0.85 (0.75, 0.98) | | |
| Flavonols, mg/d | | | | | | | |

**Supplementary Table 9. Risk of depression by quintiles of flavonoid intake levels in participants living in areas of high deprivation or with low levels of education[a] (continued)**

| Intake component | Quintile 1 (lowest) | Quintile 2 | Quintile 3 | Quintile 4 | Quintile 5 (highest) | P for trend | HR per SD (95% CI) |
|---|---|---|---|---|---|---|---|
| Intake, median (IQR) | 7.6 (4.7, 10.2) | 17.4 (15.1, 19.7) | 26.1 (24.0, 28.0) | 33.6 (31.9, 35.3) | 41.9 (39.3, 46.1) | | |
| Case/N | 498/15 729 | 409/15 728 | 441/15 728 | 449/15 728 | 403/15 728 | 0.124 | 0.96 (0.92, 1.00) |
| HR (95% CI) | 1 (reference) | 0.86 (0.76, 0.98) | 0.94 (0.83, 1.08) | 0.94 (0.83, 1.08) | 0.85 (0.74, 0.97) | | |
| Flavanones, mg/g | | | | | | | |
| Intake, median (IQR) | 0.0 (0.0, 0.1) | 0.3 (0.2, 0.4) | 0.8 (0.7, 1.1) | 2.4 (1.7, 13.1) | 35.6 (30.0, 58.4) | | |
| Case/N | 577/16 443 | 440/15 025 | 403/15 718 | 373/15 728 | 407/15 727 | **<0.001** | 0.92 (0.88, 0.96) |
| HR (95% CI) | 1 (reference) | 0.87 (0.77, 0.99) | 0.76 (0.67, 0.87) | 0.71 (0.62, 0.81) | 0.76 (0.66, 0.87) | | |
| Polymers, mg/d | | | | | | | |
| Intake, median (IQR) | 89.9 (44.2, 134.7) | 287.8 (235.8, 342.5) | 487.2 (439.7, 534.2) | 676.9 (627.9, 706.4) | 835.7 (780.1, 941.7) | | |
| Case/N | 486/15 729 | 434/15 728 | 417/15 728 | 430/15 728 | 433/15 728 | 0.314 | 0.98 (0.94, 1.03) |
| HR (95% CI) | 1 (reference) | 0.94 (0.82, 1.07) | 0.92 (0.81, 1.05) | 0.92 (0.81, 1.05) | 0.94 (0.82, 1.07) | | |
| Proanthocyanidins, mg/d[c] | | | | | | | |
| Intake, median (IQR) | 81.2 (51.4, 104.2) | 151 (136.3, 165.7) | 209.3 (194.4, 225.2) | 283.4 (261.6, 309.9) | 454.3 (388.0, 568.1) | | |
| Case/N | 497/15 729 | 458/15 728 | 405/15 728 | 414/15 728 | 426/15 728 | 0.298 | 0.99 (0.94, 1.03) |
| HR (95% CI) | 1 (reference) | 0.97 (0.85, 1.11) | 0.88 (0.76, 1.00) | 0.90 (0.78, 1.03) | 0.93 (0.81, 1.06) | | |

[a] Total number of participants, 78 641; number of participants with incident depression, 2200.

[b] Flavodiet score was calculated by summing intakes (in servings per day) of tea (black and green), red wine, apples, berries, grapes, oranges, grapefruit, sweet peppers, onions, and dark chocolate.

[c] Proanthocyanidins are also included in the polymer subclass. Model adjustment details are provided in the Methods.

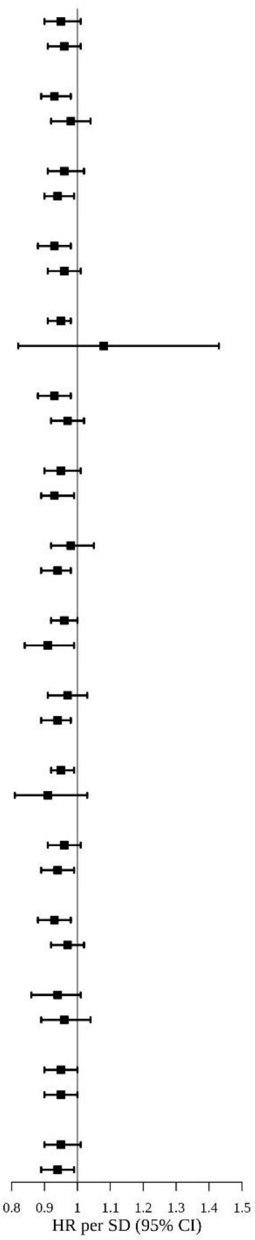


| Subgroup | Case/Person-years | Total flavonoid [median (IQR)] | HR per SD (95% CI) | P for interaction |
|---|---|---|---|---|
| Age group | | | | 0.227 |
| <60 years | 1408/580 240 | 712.8 (382.7, 1002.5) | 0.95 (0.90, 1.01) | |
| >=60 years | 1557/602 560 | 805.5 (477.8, 1058.4) | 0.96 (0.91, 1.01) | |
| Sex | | | | 0.160 |
| Female | 1894/654 193 | 744.3 (408.9, 1021.1) | 0.93 (0.89, 0.98) | |
| Male | 1071/528 607 | 782.6 (456.5, 1047.4) | 0.98 (0.92, 1.04) | |
| Socioeconomic status | | | | 0.191 |
| Less deprived | 1288/589 494 | 791.8 (461.3, 1048.7) | 0.96 (0.91, 1.02) | |
| More deprived | 1674/591 995 | 728.8 (401.7, 1015.8) | 0.94 (0.90, 0.99) | |
| Education level | | | | 0.845 |
| Lower | 1266/435 116 | 769.9 (422.4, 1032.5) | 0.93 (0.88, 0.98) | |
| Higher | 1682/744 063 | 756.0 (432.8, 1033.7) | 0.96 (0.91, 1.01) | |
| Race and ethnicity | | | | 0.543 |
| White | 2878/1 143 979 | 767.9 (435.6, 1037.1) | 0.95 (0.91, 0.98) | |
| Non-White | 73/35 065 | 544.8 (292.2, 835.0) | 1.08 (0.82, 1.43) | |
| Smoking | | | | 0.238 |
| Never | 1455/685 067 | 755.2 (424.8, 1026.7) | 0.93 (0.88, 0.98) | |
| Ever | 1498/495 219 | 768.3 (436.7, 1041.6) | 0.97 (0.92, 1.02) | |
| Physical activity | | | | 0.467 |
| Irregular | 1386/497 596 | 746.7 (410.6, 1020.1) | 0.95 (0.90, 1.01) | |
| Regular | 1463/648 467 | 772.6 (445.1, 1044.3) | 0.93 (0.89, 0.99) | |
| BMI | | | | 0.174 |
| Thin or normal | 974/476 912 | 773.9 (453.6, 1037.3) | 0.98 (0.92, 1.05) | |
| Overweight or obese | 1974/703 428 | 752.1 (412.7, 1030.6) | 0.94 (0.89, 0.98) | |
| Family history of depression | | | | 0.165 |
| No | 2311/1 031 696 | 760.4 (429.8, 1033.2) | 0.96 (0.92, 1.00) | |
| Yes | 654/151 105 | 761.2 (424.1, 1033.3) | 0.91 (0.84, 0.99) | |
| No. of medications | | | | 0.194 |
| None | 1088/610 061 | 752.9 (425.0, 1028.5) | 0.97 (0.91, 1.03) | |
| Any | 1877/572 739 | 769.3 (434.3, 1037.8) | 0.94 (0.89, 0.98) | |
| No. of chronic conditions | | | | 0.380 |
| None | 2671/1 100 751 | 758.2 (427.2, 1031.7) | 0.95 (0.92, 0.99) | |
| Any | 294/82 049 | 793.4 (458.6, 1052.0) | 0.91 (0.81, 1.03) | |
| No. of diet assessments | | | | 0.669 |
| 2 | 1291/461 462 | 761.4 (424.2, 1037.2) | 0.96 (0.91, 1.01) | |
| >3 | 1674/721 338 | 760.0 (432.1, 1030.4) | 0.94 (0.89, 0.99) | |
| hPDI | | | | 0.275 |
| 1st or 2nd quartiles | 1570/597 031 | 677.0 (351.6, 965.9) | 0.93 (0.88, 0.98) | |
| 3rd or 4th quartiles | 1395/585 770 | 847.8 (520.8, 1096) | 0.97 (0.92, 1.02) | |
| Alcohol not from red wine | | | | 0.951 |
| Under median | 653/231 410 | 765.4 (425.9, 1034.4) | 0.94 (0.86, 1.01) | |
| Above median | 704/298 521 | 767.0 (456.2, 1037.8) | 0.96 (0.89, 1.04) | |
| Energy intake | | | | 0.772 |
| Under median | 1482/590 988 | 700.1 (375.3, 988) | 0.95 (0.90, 1.00) | |
| Above median | 1483/591 812 | 823.1 (491.6, 1074.7) | 0.95 (0.90, 1.00) | |
| Fat intake | | | | 0.999 |
| Under median | 1531/591 697 | 727.9 (400.4, 1013.8) | 0.95 (0.90, 1.01) | |
| Above median | 1434/591 103 | 793.7 (464.2, 1050.6) | 0.94 (0.89, 0.99) | |

**Supplementary Figure 8. Subgroup analysis for the association of total flavonoid intake with depression risk**

[a] Model adjustment details are provided in the Methods.

**Supplementary Table 10. Risk of depression by quintiles of total flavonoid, Flavodiet Score and flavonoid subclass intake among females[a]**

| Intake component | Quintile 1 (lowest) | Quintile 2 | Quintile 3 | Quintile 4 | Quintile 5 (highest) | P for trend | HR per SD (95% CI) |
|---|---|---|---|---|---|---|---|
| Total flavonoid, mg/d | | | | | | | |
| Intake, median (IQR) | 186.3 (106.6, 261.5) | 481.4 (408.9, 550.4) | 744.3 (682.6, 806.0) | 974.0 (926.2, 1021.1) | 1214.1 (1133.8, 1343.5) | | |
| Case/N | 446/12 659 | 368/12 658 | 360/12 659 | 381/12 658 | 339/12 659 | **0.016** | 0.93 (0.89, 0.98) |
| HR (95% CI) | 1 (reference) | 0.86 (0.75, 0.99) | 0.86 (0.75, 0.99) | 0.90 (0.78, 1.03) | 0.80 (0.69, 0.93) | | |
| Flavodiet Score, points[b] | | | | | | | |
| Intake, median (IQR) | 1.1 (0.6, 1.5) | 2.6 (2.2, 2.9) | 3.8 (3.5, 4.0) | 4.7 (4.5, 5.0) | 6.0 (5.6, 6.7) | | |
| Case/N | 460/12 728 | 364/12 633 | 382/12 625 | 363/12 654 | 325/12 653 | **0.001** | 0.92 (0.88, 0.97) |
| HR (95% CI) | 1 (reference) | 0.84 (0.73, 0.96) | 0.89 (0.77, 1.02) | 0.85 (0.74, 0.98) | 0.74 (0.64, 0.86) | | |
| Anthocyanins, mg/d | | | | | | | |
| Intake, median (IQR) | 0.6 (0.0, 1.1) | 4 (2.6, 6.3) | 16.7 (12.4, 24) | 67.4 (51.6, 86.2) | 178.8 (142.4, 250.0) | | |
| Case/N | 484/13 269 | 396/12 048 | 420/12 659 | 322/12 658 | 272/12 659 | **<0.001** | 0.92 (0.88, 0.96) |
| HR (95% CI) | 1 (reference) | 0.98 (0.86, 1.13) | 0.99 (0.86, 1.13) | 0.81 (0.70, 0.94) | 0.62 (0.53, 0.73) | | |
| Flavan-3-ols, mg/d | | | | | | | |
| Intake, median (IQR) | 23.2 (13.3, 34.7) | 85.2 (66.6, 103.8) | 148.6 (134.4, 163.3) | 203.1 (190.2, 215.9) | 242.5 (232.2, 261.4) | | |
| Case/N | 434/12 659 | 384/12 658 | 353/12 659 | 377/12 658 | 346/12 659 | **0.041** | 0.95 (0.91, 1.00) |
| HR (95% CI) | 1 (reference) | 0.92 (0.80, 1.06) | 0.87 (0.76, 1.00) | 0.93 (0.80, 1.06) | 0.84 (0.73, 0.98) | | |
| Flavones, mg/d | | | | | | | |
| Intake, median (IQR) | 0.0 (0.0, 0.1) | 0.2 (0.1, 0.2) | 0.4 (0.3, 0.4) | 0.7 (0.6, 0.8) | 1.3 (1.1, 1.8) | | |
| Case/N | 445/12 659 | 346/12 658 | 351/12 659 | 351/12 658 | 401/12 659 | 0.351 | 1.03 (0.99, 1.08) |
| HR (95% CI) | 1 (reference) | 0.83 (0.72, 0.96) | 0.87 (0.75, 1.00) | 0.89 (0.77, 1.02) | 0.98 (0.85, 1.13) | | |
| Flavonols, mg/d | | | | | | | |
| Intake, median (IQR) | 7.3 (4.7, 9.9) | 17.1 (14.7, 19.5) | 25.8 (23.8, 27.8) | 33.4 (31.7, 35.1) | 41.4 (38.9, 45.4) | | |
| Case/N | 436/12 659 | 363/12 658 | 371/12 659 | 384/12 658 | 340/12 659 | 0.055 | 0.95 (0.90, 0.99) |
| HR (95% CI) | 1 (reference) | 0.87 (0.75, 1.00) | 0.91 (0.79, 1.05) | 0.94 (0.81, 1.08) | 0.82 (0.70, 0.95) | | |

**Supplementary Table 10. Risk of depression by quintiles of total flavonoid, Flavodiet Score and flavonoid subclass intake among females (continued)**

| Intake component | Quintile 1 (lowest) | Quintile 2 | Quintile 3 | Quintile 4 | Quintile 5 (highest) | P for trend | HR per SD (95% CI) |
|---|---|---|---|---|---|---|---|
| Flavanones, mg/d | | | | | | | |
| Intake, median (IQR) | 0.0 (0.0, 0.1) | 0.4 (0.3, 0.5) | 0.9 (0.7, 1.0) | 2.3 (1.6, 13.5) | 35.1 (27, 52.6) | | |
| Case/N | 492/12 772 | 396/12 576 | 308/12 628 | 336/12 658 | 362/12 659 | **<0.001** | 0.93 (0.89, 0.97) |
| HR (95% CI) | 1 (reference) | 0.88 (0.77, 1.01) | 0.68 (0.59, 0.79) | 0.74 (0.64, 0.86) | 0.79 (0.68, 0.91) | | |
| Polymers, mg/d | | | | | | | |
| Intake, median (IQR) | 92.9 (47.4, 137.4) | 288.9 (236.4, 343.5) | 493.0 (442.9, 539.2) | 680.8 (633.1, 710.8) | 839.1 (783.8, 946.1) | | |
| Case/N | 432/12 659 | 374/12 658 | 352/12 659 | 373/12 658 | 363/12 659 | 0.165 | 0.97 (0.92, 1.01) |
| HR (95% CI) | 1 (reference) | 0.90 (0.79, 1.04) | 0.87 (0.76, 1.01) | 0.91 (0.79, 1.05) | 0.89 (0.77, 1.03) | | |
| Proanthocyanidins, mg/d[c] | | | | | | | |
| Intake, median (IQR) | 84.0 (54.3, 106.9) | 154.5 (139.8, 168.8) | 211.9 (197.4, 227.2) | 284.5 (263.4, 310.0) | 450.7 (387, 565.2) | | |
| Case/N | 437/12 659 | 391/12 658 | 337/12 659 | 365/12 658 | 364/12 659 | 0.287 | 0.98 (0.93, 1.03) |
| HR (95% CI) | 1 (reference) | 0.95 (0.83, 1.10) | 0.85 (0.73, 0.98) | 0.92 (0.80, 1.07) | 0.91 (0.78, 1.05) | | |

[a] Total number of participants, 63 293; number of participants with incident depression, 1894.

[b] Flavodiet score was calculated by summing intakes (in servings per day) of tea (black and green), red wine, apples, berries, grapes, oranges, grapefruit, sweet peppers, onions, and dark chocolate.

[c] Proanthocyanidins are also included in the polymer subclass. Model adjustment details are provided in the Methods

**Supplementary Table 11. Risk of depression by quintiles of total flavonoid, Flavodiet Score and flavonoid subclass intake among males[a]**

| Intake component | Quintile 1 (lowest) | Quintile 2 | Quintile 3 | Quintile 4 | Quintile 5 (highest) | P for trend | HR per SD (95% CI) |
|---|---|---|---|---|---|---|---|
| Total flavonoid, mg/d | | | | | | | |
| Intake, median (IQR) | 205.9 (110.3, 297.1) | 526.4 (456.5, 595.6) | 782.6 (720.9, 842) | 998.5 (951.2, 1047.4) | 1263.2 (1171.2, 1408.4) | 0.493 | 0.98 (0.92, 1.04) |
| Case/N | 226/10 311 | 192/10 311 | 231/10 311 | 225/10 311 | 197/10 311 | | |
| HR (95% CI) | 1 (reference) | 0.89 (0.74, 1.08) | 1.06 (0.88, 1.27) | 0.99 (0.82, 1.2) | 0.87 (0.72, 1.07) | | |
| Flavodiet Score, points[b] | | | | | | | |
| Intake, median (IQR) | 1.2 (0.6, 1.6) | 2.7 (2.4, 3.0) | 3.8 (3.5, 4.0) | 4.8 (4.5, 5.0) | 6.2 (5.8, 7.0) | 0.341 | 0.96 (0.90, 1.02) |
| Case/N | 229/11 031 | 201/9601 | 235/10 408 | 216/10 207 | 190/10 308 | | |
| HR (95% CI) | 1 (reference) | 1.05 (0.86, 1.27) | 1.1 (0.92, 1.33) | 1.04 (0.86, 1.26) | 0.88 (0.72, 1.09) | | |
| Anthocyanins, mg/d | | | | | | | |
| Intake, median (IQR) | 0.0 (0.0, 0.6) | 2.1 (1.6, 3.0) | 12.9 (7.8, 24.8) | 94.5 (77.2, 120.5) | 247.1 (189.8, 337.3) | 0.019 | 0.89 (0.85, 0.95) |
| Case/N | 250/10 324 | 259/10 298 | 219/10 311 | 173/10 556 | 170/10 066 | | |
| HR (95% CI) | 1 (reference) | 1.13 (0.95, 1.35) | 0.96 (0.80, 1.16) | 0.78 (0.64, 0.96) | 0.72 (0.59, 0.89) | | |
| Flavan-3-ols, mg/d | | | | | | | |
| Intake, median (IQR) | 30.5 (16.9, 44.6) | 95.7 (79.3, 112.5) | 154.8 (141.5, 168.6) | 208.6 (195.3, 219.6) | 249.0 (237.3, 270.0) | 0.906 | 1.00 (0.94, 1.06) |
| Case/N | 217/10 311 | 202/10 311 | 221/10 311 | 222/10 311 | 209/10 311 | | |
| HR (95% CI) | 1 (reference) | 0.97 (0.80, 1.18) | 1.04 (0.86, 1.26) | 1.02 (0.84, 1.23) | 0.96 (0.79, 1.17) | | |
| Flavones, mg/d | | | | | | | |
| Intake, median (IQR) | 0.0 (0.0, 0.1) | 0.1 (0.1, 0.2) | 0.3 (0.2, 0.3) | 0.5 (0.4, 0.6) | 1.1 (0.8, 1.5) | 0.185 | 0.96 (0.90, 1.03) |
| Case/N | 241/10 311 | 231/10 361 | 185/10 261 | 215/10 311 | 199/10 311 | | |
| HR (95% CI) | 1 (reference) | 1.01 (0.84, 1.22) | 0.83 (0.68, 1.01) | 0.97 (0.80, 1.17) | 0.87 (0.71, 1.06) | | |
| Flavonols, mg/d | | | | | | | |
| Intake, median (IQR) | 8.5 (5.4, 11.2) | 18.5 (16.1, 20.7) | 26.9 (24.9, 28.9) | 34.3 (32.6, 36.1) | 42.7 (40.0, 46.8) | 0.588 | 1.01 (0.95, 1.08) |
| Case/N | 223/10 311 | 187/10 311 | 208/10 311 | 231/10 311 | 222/10 311 | | |
| HR (95% CI) | 1 (reference) | 0.85 (0.70, 1.04) | 0.95 (0.78, 1.15) | 1.03 (0.86, 1.25) | 0.98 (0.80, 1.19) | | |

**Supplementary Table 11. Risk of depression by quintiles of total flavonoid, Flavodiet Score and flavonoid subclass intake among males (continued)**

| | | | | | | | |
|---|---|---|---|---|---|---|---|
| Flavanones, mg/d | | | | | | | |
| Intake, median (IQR) | 0.0 (0.0, 0.0) | 0.3 (0.2, 0.4) | 1.0 (0.8, 1.2) | 2.9 (1.9, 16.7) | 40.9 (34.1, 68.1) | | |
| Case/N | 248/10 312 | 225/10 624 | 191/9997 | 188/10 351 | 219/10 271 | 0.101 | 0.96 (0.90, 1.02) |
| HR (95% CI) | 1 (reference) | 0.94 (0.78, 1.13) | 0.86 (0.71, 1.04) | 0.79 (0.65, 0.96) | 0.92 (0.76, 1.11) | | |
| Polymers, mg/d | | | | | | | |
| Intake, median (IQR) | 95.3 (46.2, 146.6) | 310.2 (256.7, 361) | 503.4 (455.8, 546.8) | 682.8 (638, 712.1) | 846.5 (787.6, 959.7) | | |
| Case/N | 217/10 311 | 192/10 311 | 226/10 311 | 223/10 311 | 213/10 311 | 0.603 | 1.03 (0.96, 1.09) |
| HR (95% CI) | 1 (reference) | 0.94 (0.77, 1.14) | 1.09 (0.91, 1.32) | 1.03 (0.85, 1.25) | 1.01 (0.83, 1.23) | | |
| Proanthocyanidins, mg/d[c] | | | | | | | |
| Intake, median (IQR) | 84.6 (54.0, 107.7) | 154.8 (140.0, 169.9) | 214.3 (199.1, 230.7) | 291.0 (268.2, 319.7) | 469.8 (402.3, 590.0) | | |
| Case/N | 225/10 311 | 216/10 311 | 218/10 311 | 203/10 311 | 209/10 311 | 0.738 | 0.99 (0.92, 1.05) |
| HR (95% CI) | 1 (reference) | 0.99 (0.82, 1.19) | 1.01 (0.84, 1.23) | 0.95 (0.78, 1.16) | 0.97 (0.80, 1.19) | | |

[a] Total number of participants, 51 555; number of participants with incident depression, 1071.

[b] Flavodiet score was calculated by summing intakes (in servings per day) of tea (black and green), red wine, apples, berries, grapes, oranges, grapefruit, sweet peppers, onions, and dark chocolate.

[c] Proanthocyanidins are also included in the polymer subclass. Model adjustment details are provided in the Methods

**Supplementary Table 12. Association of total flavonoid intake with potential mediators as physical measures, biochemical indices, and history of chronic diseases[a]**

| | Beta per SD | P value |
|---|---|---|
| Hand grip strength | 1.09E-01 | **2.83E-07** |
| Muscle Mass Index | 4.13E-02 | **4.18E-08** |
| Body fat percentage | -1.51E-02 | 2.21E-01 |
| Whole body fat mass | -7.83E-02 | **5.31E-07** |
| Whole body fat free mass | -6.39E-02 | **2.91E-05** |
| Whole body water mass | -4.78E-02 | **2.17E-05** |
| Basal metabolic rate | -9.35E+00 | **9.50E-07** |
| Leg fat percentage (right) | -7.78E-03 | 4.69E-01 |
| Leg fat mass (right) | -1.43E-02 | **6.83E-08** |
| Leg fat free mass (right) | -1.53E-02 | **6.40E-08** |
| Leg predicted mass (right) | -1.42E-02 | **8.42E-08** |
| Leg fat percentage (left) | -2.86E-03 | 7.75E-01 |
| Leg fat mass (left) | -1.40E-02 | **5.41E-08** |
| Leg fat free mass (left) | -1.61E-02 | **3.67E-09** |
| Leg predicted mass (left) | -1.52E-02 | **2.97E-09** |
| Arm fat percentage (right) | -3.55E-02 | **1.07E-02** |
| Arm fat mass (right) | -7.38E-03 | **3.04E-12** |
| Arm fat free mass (right) | -4.76E-03 | **7.82E-06** |
| Arm predicted mass (right) | -4.65E-03 | **3.58E-06** |
| Arm fat percentage (left) | -5.03E-02 | **2.47E-04** |
| Arm fat mass (left) | -8.80E-03 | **1.76E-13** |
| Arm fat free mass (left) | -3.89E-03 | **5.19E-04** |
| Arm predicted mass (left) | -3.62E-03 | **9.05E-04** |
| Trunk fat percentage | -1.39E-02 | 3.75E-01 |
| Trunk fat mass | -3.31E-02 | **5.34E-04** |
| Trunk fat free mass | -2.39E-02 | **5.17E-03** |
| Trunk predicted mass | -2.34E-02 | **4.31E-03** |
| Alanine aminotransferase | -1.29E-01 | **1.01E-03** |
| Albumin | -8.70E-03 | 3.05E-01 |
| Alkaline phosphatase | 4.16E-01 | **2.47E-08** |
| Apolipoprotein A | 9.47E-03 | **<2.00E-16** |
| Apolipoprotein B | -5.69E-03 | **6.18E-15** |
| Aspartate aminotransferase | -7.93E-03 | 7.89E-01 |
| C reactive protein | -8.74E-03 | 4.75E-01 |
| Calcium | 1.73E-03 | **2.58E-08** |
| Cholesterol | -6.59E-03 | 5.47E-02 |
| Creatinine | 5.12E-01 | **<2.00E-16** |
| Cystatin C | -3.19E-03 | **3.10E-13** |
| Creatine to Cystatin-C ratio | 8.44E-01 | **<2.00E-16** |
| Direct bilirubin | 1.02E-02 | **1.57E-04** |
| Gamma glutamyltransferase | 6.40E-01 | **8.91E-10** |
| Glucose | -1.19E-02 | **7.82E-04** |
| Glycated haemoglobin | -5.42E-02 | **1.95E-03** |

**Supplementary Table 12. Association of total flavonoid intake with potential mediators as physical measures, biochemical indices, and history of chronic diseases[a] (continued)**

| | Beta per SD | P value |
|---|---|---|
| HDL-cholesterol | 1.59E-02 | **<2.00E-16** |
| IGF-1 | 7.74E-02 | **6.94E-06** |
| LDL direct | -1.38E-02 | **1.85E-07** |
| Lipoprotein A | 4.59E-02 | 7.94E-01 |
| Oestradiol | 2.52E+00 | 4.69E-01 |
| Phosphate | 1.07E-02 | **<2.00E-16** |
| Rheumatoid factor | -4.05E-01 | 6.57E-02 |
| SHBG | -1.83E-01 | **2.43E-02** |
| Testosterone | -7.17E-03 | 4.03E-01 |
| Total bilirubin | 5.25E-02 | **1.74E-04** |
| Total protein | 5.75E-02 | **1.64E-05** |
| Triglycerides | -3.01E-02 | **<2.00E-16** |
| Urate | 4.87E-01 | **1.27E-02** |
| Urea | -3.72E-02 | **<2.00E-16** |
| Vitamin D | 5.69E-01 | **<2.00E-16** |
| Cancer | -5.75E-03 | 6.39E-01 |
| Chronic obstructive pulmonary disease | -2.70E-02 | 2.34E-01 |
| Main diseases of circulatory system | -9.96E-03 | 5.73E-01 |
| Main diseases of nervous system | -8.53E-02 | 2.64E-01 |
| Main diseases of endorine system | -8.04E-02 | **4.04E-08** |

**Supplementary Table 13. Participant characteristics according to the availability of two brain MRI scans**

| Characteristics | Value, median (IQR) | |
|---|---|---|
| | Without twice brain MRI scans (n = 112 728) | With twice brain MRI scans (n = 2120) |
| **Dietary** | | |
| Total flavonoid, mg/d | 760.2 (429.1, 1033.1) | 779.1 (429.5, 1038.6) |
| Anthocyanins, mg/d | 15.3 (1.9, 102.5) | 15.7 (1.7, 100.1) |
| Flavan-3-ols, mg/d | 151.5 (72.3, 217.5) | 152.5 (76.3, 217.1) |
| Flavones, mg/d | 0.3 (0.1, 0.7) | 0.3 (0.1, 0.7) |
| Flavonols, mg/d | 26.3 (15.4, 35.5) | 26.7 (15.6, 35.4) |
| Flavanones, mg/d | 0.9 (0.3, 14.5) | 0.8 (0.2, 11.6) |
| Polymers, mg/d | 497.4 (245.6, 711.1) | 508.6 (251.5, 714.9) |
| Proanthocyanidins, mg/d[a] | 213.0 (140.0, 314.0) | 212.4 (138.0, 319.0) |
| Flavodiet score (tea capped at 4 cups), servings/d[b] | 3.8 (2.2, 5.0) | 3.8 (2.2, 5.0) |
| Tea | 2.0 (0.7, 3.5) | 2.2 (0.7, 3.5) |
| Red wine | 0.0 (0.0, 0.8) | 0.0 (0.0, 0.8) |
| Apples | 0.2 (0.0, 0.7) | 0.2 (0.0, 0.7) |
| Berries | 0.0 (0.0, 0.2) | 0.0 (0.0, 0.2) |
| Grapes | 0.0 (0.0, 0.2) | 0.0 (0.0, 0.2) |
| Grapefruit | 0.0 (0.0, 0.0) | 0.0 (0.0, 0.0) |
| Oranges | 0.0 (0.0, 0.0) | 0.0 (0.0, 0.0) |
| Peppers | 0.0 (0.0, 0.1) | 0.0 (0.0, 0.1) |
| Onions | 0.0 (0.0, 0.2) | 0.1 (0.0, 0.2) |
| Dark chocolate | 0.0 (0.0, 0.0) | 0.0 (0.0, 0.0) |
| Energy, kcal/d | 2008.7 (1718.0, 2330.3) | 2030.1 (1754.1, 2348.9) |
| Fat, g/d | 70.3 (56.1, 86.4) | 71.2 (57.0, 87.2) |
| Alcohol not from red wine, g/d | 5.1 (1.7, 12.0) | 6.9 (1.7, 13.7) |
| hPDI score | 60.8 (57.7, 63.8) | 60.5 (57.5, 63.7) |
| Dietary assessment completion, d | 3.0 (2.0, 4.0) | 3.0 (2.0, 4.0) |
| **Demographic** | | |
| Age, y | 60.5 (53.2, 65.4) | 56.0 (49.9, 62.1) |
| Duration of follow-up, y | 10.5 (10.4, 10.9) | 10.5 (10.4, 10.9) |
| Physical activity, No. (%) | | |
| Irregular | 47 663 (42.3) | 820 (38.7) |
| Regular | 61 535 (54.6) | 1242 (58.6) |
| Unknown | 3530 (3.1) | 58 (2.7) |
| BMI, kg/m$^2$ | 26.0 (23.5, 28.9) | 25.7 (23.3, 28.5) |
| Sex, No. (%) | | |
| Female | 62 166 (55.1) | 1127 (53.2) |
| Male | 50 562 (44.9) | 993 (46.8) |
| Race and ethnicity, No. (%) | | |
| Asian | 990 (0.9) | 16 (0.8) |

**Supplementary Table 13. Participant characteristics according to the availability of two brain MRI scans (continued)**

| Characteristics | Value, median (IQR) Without twice brain MRI scans (n = 112 728) | With twice brain MRI scans (n = 2120) |
|---|---|---|
| Black | 824 (0.7) | 17 (0.8) |
| Chinese | 290 (0.3) | 7 (0.3) |
| White | 109 066 (96.8) | 2061 (97.2) |
| Multiracial | 570 (0.5) | 8 (0.4) |
| Other[c] | 618 (0.5) | 8 (0.4) |
| Unknown | 370 (0.3) | 3 (0.1) |
| Townsend deprivation index, No. (%) | | |
| Low | 22 496 (20.0) | 455 (21.5) |
| Moderate | 67 528 (59.9) | 1302 (61.4) |
| High | 22 583 (20.0) | 359 (16.9) |
| Unknown | 121 (0.1) | 4 (0.2) |
| Education level, No. (%) | | |
| Lower secondary | 15 360 (13.6) | 247 (11.7) |
| Upper secondary | 7015 (6.2) | 119 (5.6) |
| Vocational | 11 993 (10.6) | 260 (12.3) |
| Higher | 70 675 (62.7) | 1424 (67.2) |
| Other | 7330 (6.5) | 68 (3.2) |
| Unknown | 355 (0.3) | 2 (0.1) |
| Smoking, No. (%) | | |
| Ever | 47 762 (42.4) | 765 (36.1) |
| Never | 64 722 (57.4) | 1352 (63.8) |
| Unknown | 244 (0.2) | 3 (0.1) |
| Sleep duration, No. (%) | | |
| >8 | 6500 (5.8) | 124 (5.8) |
| 7-8 | 82 225 (72.9) | 1561 (73.6) |
| ≤6 | 23 767 (21.1) | 432 (20.4) |
| Unknown | 236 (0.2) | 3 (0.1) |
| Family history of depression, No. (%) | | |
| Yes | 14 452 (12.8) | 273 (12.9) |
| No | 98 276 (87.2) | 1847 (87.1) |
| No. of medications taken, No. (%) | | |
| 0 | 57 545 (51.0) | 1220 (57.5) |
| 1 | 34 648 (30.7) | 647 (30.5) |
| 2 | 12 484 (11.1) | 176 (8.3) |
| 3 | 6607 (5.9) | 69 (3.3) |
| >4 | 1444 (1.3) | 8 (0.4) |
| No. of chronic conditions, No. (%) | | |
| 0 | 104 433 (92.6) | 2026 (95.6) |
| 1 | 8028 (7.1) | 91 (4.3) |
| >2 | 267 (0.2) | 3 (0.1) |

Abbreviations: BMI, body mass index; hPDI, healthful plant-based diet index.

[a] Proanthocyanidins are also included in the polymer subclass.

[b] Flavodiet score was calculated by summing intakes (in servings per day) of tea (black and green), red wine, apples, berries, grapes, oranges, grapefruit, sweet peppers, onions, and dark chocolate.

[c] Other race and ethnicity was self-selected and includes no additional information.

**Supplementary Table 14. Associations of total flavonoid intake (per SD) with brain structural changes[a]**

| Region | Left | | Right | |
|---|---|---|---|---|
| | Beta | *P value* | Beta | *P value* |
| **Volume of brain** | | | | |
| Total gray matter | -0.006 | 0.412 | \ | \ |
| Total white matter | 0.006 | 0.537 | \ | \ |
| White matter hyperintensities | 0.018 | 0.180 | \ | \ |
| Total brain (gray + white) | 0.000 | 0.994 | \ | \ |
| **Volume of cortex** | | | | |
| Bankssts | -0.007 | 0.425 | -0.013 | 0.156 |
| Caudalanteriorcingulate | -0.026 | **0.021** | 0.009 | 0.447 |
| Caudalmiddlefrontal | -0.018 | **0.041** | -0.028 | **0.003** |
| Cuneus | -0.013 | 0.178 | 0.004 | 0.688 |
| Entorhinal | 0.010 | 0.542 | 0.004 | 0.795 |
| Fusiform | -0.006 | 0.564 | -0.001 | 0.891 |
| Inferiorparietal | -0.009 | 0.303 | -0.007 | 0.413 |
| Inferiortemporal | -0.017 | 0.059 | 0.006 | 0.519 |
| Isthmuscingulate | -0.011 | 0.246 | -0.01 | 0.297 |
| Lateraloccipital | -0.015 | 0.170 | -0.006 | 0.544 |
| Lateralorbitofrontal | -0.010 | 0.414 | -0.004 | 0.802 |
| Lingual | -0.011 | 0.297 | -0.005 | 0.618 |
| Medialorbitofrontal | -0.007 | 0.649 | -0.001 | 0.917 |
| Middletemporal | -0.008 | 0.442 | 0.005 | 0.621 |
| Parahippocampal | -0.015 | 0.206 | -0.005 | 0.706 |
| Paracentral | -0.009 | 0.488 | -0.012 | 0.277 |
| Parsopercularis | -0.009 | 0.237 | -0.01 | 0.293 |
| Parsorbitalis | -0.019 | 0.147 | 0.021 | 0.073 |
| Parstriangularis | -0.004 | 0.597 | 0.007 | 0.379 |
| Pericalcarine | -0.012 | 0.182 | 0.000 | 0.992 |
| Postcentral | -0.013 | 0.278 | -0.014 | 0.268 |
| Posteriorcingulate | -0.014 | 0.182 | 0.005 | 0.659 |
| Precentral | -0.016 | 0.213 | -0.030 | **0.020** |
| Precuneus | -0.015 | 0.136 | -0.008 | 0.441 |
| Rostralanteriorcingulate | -0.003 | 0.782 | -0.004 | 0.693 |
| Rostralmiddlefrontal | -0.008 | 0.455 | 0.013 | 0.186 |
| Superiorfrontal | -0.015 | 0.173 | -0.005 | 0.631 |
| Superiorparietal | -0.015 | 0.179 | -0.008 | 0.488 |
| Superiortemporal | -0.021 | **0.046** | -0.001 | 0.936 |
| Supramarginal | -0.017 | **0.049** | -0.006 | 0.528 |
| Frontalpole | -0.006 | 0.729 | -0.017 | 0.350 |

**Supplementary Table 14. Associations of total flavonoid intake (per SD) with brain structural changes[a] (continued)**

| Region | Left | | Right | |
|---|---|---|---|---|
| | Beta | *P value* | Beta | *P value* |
| Transversetemporal | -0.001 | 0.954 | 0.013 | 0.278 |
| Insula | -0.010 | 0.497 | 0.012 | 0.490 |
| **FA of white matter tracts** | | | | |
| Acoustic Radiation | 0.010 | 0.324 | 0.010 | 0.401 |
| Anterior Thalamic Radiation | 0.001 | 0.871 | 0.005 | 0.511 |
| Cingulate Gyrus Part of Cingulum | 0.002 | 0.848 | 0.016 | 0.088 |
| Parahippocampal Part of Cingulum | 0.021 | 0.177 | 0.014 | 0.346 |
| Corticospinal Tract | 0.003 | 0.735 | 0.007 | 0.465 |
| Inferior Fronto-Occipital Fasciculus | 0.007 | 0.346 | 0.006 | 0.430 |
| Inferior Longitudinal Fasciculus | 0.001 | 0.839 | 0.004 | 0.357 |
| Medial Lemniscus | -0.011 | 0.386 | -0.011 | 0.378 |
| Posterior Thalamic Radiation | 0.001 | 0.950 | 0.012 | 0.160 |
| Superior Longitudinal Fasciculus | -0.003 | 0.534 | 0.002 | 0.677 |
| Superior Thalamic Radiation | -0.001 | 0.921 | -0.001 | 0.823 |
| Uncinate Fasciculus | 0.006 | 0.518 | 0.012 | 0.141 |
| Forceps Major | 0.001 | 0.863 | \ | \ |
| Forceps Minor | 0.003 | 0.532 | \ | \ |
| Middle Cerebellar Peduncle | 0.010 | 0.432 | \ | \ |
| **MD of white matter tracts** | | | | |
| Acoustic Radiation | 0.017 | 0.116 | 0.007 | 0.515 |
| Anterior Thalamic Radiation | 0.021 | **0.018** | 0.015 | 0.113 |
| Cingulate Gyrus Part of Cingulum | 0.010 | 0.253 | 0.006 | 0.483 |
| Parahippocampal Part of Cingulum | 0.013 | 0.314 | 0.018 | 0.167 |
| Corticospinal Tract | 0.013 | 0.183 | 0.003 | 0.714 |
| Inferior Fronto-Occipital Fasciculus | 0.012 | 0.176 | 0.012 | 0.190 |
| Inferior Longitudinal Fasciculus | 0.009 | 0.189 | 0.006 | 0.398 |
| Medial Lemniscus | 0.007 | 0.537 | -0.008 | 0.420 |
| Posterior Thalamic Radiation | 0.010 | 0.303 | 0.015 | 0.130 |
| Superior Longitudinal Fasciculus | 0.006 | 0.221 | 0.005 | 0.347 |
| Superior Thalamic Radiation | 0.003 | 0.566 | 0.005 | 0.374 |
| Uncinate Fasciculus | 0.010 | 0.318 | 0.003 | 0.680 |
| Forceps Major | 0.007 | 0.420 | \ | \ |
| Forceps Minor | 0.003 | 0.658 | \ | \ |
| Middle Cerebellar Peduncle | 0.025 | 0.084 | \ | \ |

[a] Total number of participants, 2120. Model adjustment details are provided in the

Methods.

**Supplementary Table 15. Associations of Flavodiet Score (per serving) with brain structural changes**

| | Left | | Right | |
|---|---|---|---|---|
| Region | Beta | *P value* | Beta | *P value* |
| **Volume of brain** | | | | |
| Total gray matter | -0.005 | 0.226 | \ | \ |
| Total white matter | 0.013 | **0.020** | \ | \ |
| White matter hyperintensities | 0.013 | 0.085 | \ | \ |
| Total brain (gray + white) | 0.004 | 0.214 | \ | \ |
| **Volume of cortex** | | | | |
| Bankssts | -0.004 | 0.325 | -0.008 | 0.120 |
| Caudalanteriorcingulate | -0.018 | **0.003** | 0.006 | 0.305 |
| Caudalmiddlefrontal | -0.007 | 0.141 | -0.012 | **0.015** |
| Cuneus | -0.002 | 0.649 | 0.005 | 0.392 |
| Entorhinal | 0.005 | 0.544 | 0.009 | 0.270 |
| Fusiform | -0.003 | 0.564 | 0.003 | 0.586 |
| Inferiorparietal | -0.005 | 0.262 | -0.006 | 0.220 |
| Inferiortemporal | -0.012 | **0.014** | 0.002 | 0.624 |
| Isthmuscingulate | -0.001 | 0.872 | -0.002 | 0.777 |
| Lateraloccipital | -0.005 | 0.419 | -0.001 | 0.844 |
| Lateralorbitofrontal | -0.01 | 0.148 | 0.001 | 0.936 |
| Lingual | -0.003 | 0.525 | 0.003 | 0.555 |
| Medialorbitofrontal | 0.004 | 0.606 | 0.004 | 0.619 |
| Middletemporal | -0.006 | 0.314 | 0.002 | 0.712 |
| Parahippocampal | -0.006 | 0.323 | -0.001 | 0.881 |
| Paracentral | -0.006 | 0.424 | -0.006 | 0.348 |
| Parsopercularis | -0.005 | 0.239 | -0.003 | 0.529 |
| Parsorbitalis | -0.005 | 0.498 | 0.012 | 0.053 |
| Parstriangularis | -0.003 | 0.504 | 0.001 | 0.841 |
| Pericalcarine | -0.005 | 0.282 | 0.003 | 0.601 |
| Postcentral | -0.006 | 0.336 | -0.007 | 0.283 |
| Posteriorcingulate | -0.009 | 0.133 | 0.005 | 0.454 |
| Precentral | -0.009 | 0.177 | -0.013 | 0.058 |
| Precuneus | -0.008 | 0.130 | -0.002 | 0.671 |
| Rostralanteriorcingulate | -0.006 | 0.327 | -0.003 | 0.509 |
| Rostralmiddlefrontal | -0.004 | 0.488 | 0.007 | 0.219 |
| Superiorfrontal | -0.006 | 0.337 | -0.002 | 0.714 |
| Superiorparietal | -0.008 | 0.179 | -0.006 | 0.328 |
| Superiortemporal | -0.011 | 0.057 | 0.000 | 0.952 |
| Supramarginal | -0.011 | **0.023** | -0.008 | 0.137 |
| Frontalpole | 0.002 | 0.843 | -0.002 | 0.806 |

**Supplementary Table 15. Associations of Flavodiet Score (per serving) with brain structural changes (continued)**

| | Left | | Right | |
|---|---|---|---|---|
| **Region** | **Beta** | **P value** | **Beta** | **P value** |
| Transversetemporal | 0.001 | 0.892 | 0.003 | 0.609 |
| Insula | -0.011 | 0.168 | -0.001 | 0.904 |
| **FA of white matter tracts** | | | | |
| Acoustic Radiation | 0.006 | 0.268 | 0.006 | 0.392 |
| Anterior Thalamic Radiation | -0.001 | 0.847 | 0.000 | 0.923 |
| Cingulate Gyrus Part of Cingulum | 0.001 | 0.899 | 0.006 | 0.241 |
| Parahippocampal Part of Cingulum | 0.006 | 0.455 | 0.007 | 0.384 |
| Corticospinal Tract | -0.002 | 0.669 | 0.002 | 0.689 |
| Inferior Fronto-Occipital Fasciculus | 0.003 | 0.511 | 0.001 | 0.738 |
| Inferior Longitudinal Fasciculus | 0.001 | 0.811 | 0.002 | 0.564 |
| Medial Lemniscus | -0.001 | 0.829 | -0.001 | 0.860 |
| Posterior Thalamic Radiation | 0 | 0.945 | 0.005 | 0.298 |
| Superior Longitudinal Fasciculus | 0.001 | 0.714 | 0.003 | 0.275 |
| Superior Thalamic Radiation | 0.002 | 0.539 | -0.001 | 0.829 |
| Uncinate Fasciculus | 0.004 | 0.432 | 0.005 | 0.279 |
| Forceps Major | 0.002 | 0.552 | \ | \ |
| Forceps Minor | 0.001 | 0.607 | \ | \ |
| Middle Cerebellar Peduncle | 0.001 | 0.898 | \ | \ |
| **MD of white matter tracts** | | | | |
| Acoustic Radiation | 0.003 | 0.611 | 0.002 | 0.798 |
| Anterior Thalamic Radiation | 0.008 | 0.097 | 0.005 | 0.301 |
| Cingulate Gyrus Part of Cingulum | 0.003 | 0.544 | 0.002 | 0.590 |
| Parahippocampal Part of Cingulum | 0.008 | 0.241 | 0.004 | 0.606 |
| Corticospinal Tract | 0.004 | 0.478 | -0.002 | 0.655 |
| Inferior Fronto-Occipital Fasciculus | 0 | 0.982 | 0.002 | 0.721 |
| Inferior Longitudinal Fasciculus | 0 | 0.894 | 0.001 | 0.888 |
| Medial Lemniscus | 0.001 | 0.854 | -0.01 | 0.076 |
| Posterior Thalamic Radiation | -0.001 | 0.853 | 0.004 | 0.423 |
| Superior Longitudinal Fasciculus | 0.001 | 0.780 | 0.001 | 0.848 |
| Superior Thalamic Radiation | 0 | 0.923 | 0.002 | 0.401 |
| Uncinate Fasciculus | 0.002 | 0.760 | -0.002 | 0.654 |
| Forceps Major | 0 | 0.938 | \ | \ |
| Forceps Minor | -0.001 | 0.867 | \ | \ |
| Middle Cerebellar Peduncle | 0.008 | 0.305 | \ | \ |

[a] Total number of participants, 2120. Model adjustment details are provided in the

Methods.

[12] WineGB 2018. International Cool Climate Wine Symposium (ICCWS) & WineGB Yield Survey 2018 [M].

[13] Di Vita G, Zanchini R, Spina D, et al. Simply red? The effects of distinct colours and sustainable production methods on the consumers’ preferences for healthier sweet peppers[J]. Heliyon, 2024, 10(8).

[14] Griffiths G, Trueman L, Crowther T, et al. Onions—a global benefit to health[J]. Phytotherapy Research: An International Journal Devoted to Pharmacological and Toxicological Evaluation of Natural Product Derivatives, 2002, 16(7): 603-615.

[15] Weber A M, Mawodza T, Sarkar B, et al. Assessment of potentially toxic trace element contamination in urban allotment soils and their uptake by onions: A preliminary case study from Sheffield, England[J]. Ecotoxicology and Environmental Safety, 2019, 170: 156-165.

[16] Bhagwat S, Haytowitz D B, Wasswa-Kintu S I, et al. USDA develops a database for flavonoids to assess dietary intakes[J]. Procedia Food Science, 2013, 2: 81-86.

[17] Crawley H. Food portion sizes[C]//London: HMSO:Ministry of Agriculture FaF,1988

[18] Galante J, Adamska L, Young A, et al. The acceptability of repeat Internet-based hybrid diet assessment of previous 24-h dietary intake: administration of the Oxford WebQ in UK Biobank[J]. British Journal of Nutrition, 2016, 115(4): 681-686.

[19] Brown R, Gray A R, Chua M G, et al. Is a handful an effective way to guide nut

recommendations?[J]. International journal of environmental research and public health, 2021, 18(15): 7812.
[20] Lang A 2019. 7 Interesting Types of Bean Sprouts [M]. healthline.
[21] Porter T B D A S 2021. What is a portion of food? [M].
[22] Thompson A S, Tresserra-Rimbau A, Karavasiloglou N, et al. Association of Healthful Plant-based Diet Adherence With Risk of Mortality and Major Chronic Diseases Among Adults in the UK[J]. JAMA Netw Open, 2023, 6(3): e234714.
